# Bayesian Mendelian Randomization Analysis for Latent Exposures Leveraging GWAS Summary Statistics for Traits Co-Regulated by the Exposures

**DOI:** 10.1101/2024.11.25.24317939

**Authors:** Yue Yu, Andrew Lakkis, Bingxin Zhao, Jin Jin

## Abstract

Mendelian Randomization analysis is a popular method to infer causal relationships between exposures and outcomes, utilizing data from genome-wide association studies (GWAS) to overcome limitations of observational research by treating genetic variants as instrumental variables. This study focuses on a specific problem setting, where causal signals may exist among a series of correlated traits, but the exposures of interest, such as biological functions or lower-dimensional latent factors that regulate the observable traits, are not directly observable. We propose a Bayesian Mendelian randomization analysis framework that allows joint analysis of the causal effects of multiple latent exposures on a disease outcome leveraging GWAS summary-level association statistics for traits co-regulated by the exposures. We conduct simulation studies to show the validity and superiority of the method in terms of type I error control and power due to a more flexible modeling framework and a more stable algorithm compared to an alternative approach and traditional single- and multi-exposure analysis approaches not specifically designed for the problem. We have also applied the method to reveal evidence of the causal effects of psychiatric factors, including compulsive, psychotic, neurodevelopmental, and internalizing factors, on neurodegenerative, autoimmune, digestive, and cardiometabolic diseases.

## 1 Introduction

Mendelian randomization (MR) is a widely implemented tool for causal inference utilizing observational data from genome-wide association studies (GWAS) when performing a randomized controlled trial is not practical (12; 29). As a type of instrumental variable (IV) analysis, MR employs genetic variants, typically single nucleotide polymorphisms (SNPs), as IVs to infer causal relationships between exposures and outcomes in the presence of unmeasured confounding. As the availability of summary data from GWAS continues to increase, replacing the reliance on individual-level data, recent studies have focused on exploiting the GWAS summary-level data to perform MR (6; 40), providing valuable insight into the mechanisms across a wide spectrum of human traits and diseases. Extensive methodological advancements have been made in relaxing the classic MR assumptions and proposing more robust analysis approaches. Traditionally, three assumptions are required for an IV to be valid: it must be truly associated with the exposure, it only affects the outcome through the exposure (no horizontal pleiotropy), and it is not associated with confounders of the association between the exposure and the outcome (“exchange-ability”) (6; 15; 33). These assumptions, however, are over-stringent and often hard to verify. The widely implemented two-sample MR (39), with outcome and exposure data coming from two non-overlapping GWAS, can mitigate the violation of the exchangeability assumption to a reasonable degree. Endeavors toward developing robust and powerful methods include accounting for uncorrelated and correlated horizontal pleiotropy (37; 9; 41), correcting for weak instrumental bias(45; 10; 32), and adjusting for population stratification and batch effects (3; 10; 19), among others. Recent work also started to explore MR frameworks for high-dimensional exposures in the contexts of multi-omic and medical imaging (7; 24; 34; 26) research.

While current MR analyses focus on well-defined exposures for which GWAS can be directly conducted, the underlying causal factors may be some lower-dimensional latent features that regulate the observed biomarker traits. Such a latent exposure setting is quite common: we often encounter scenarios where the latent exposures of interest may not be directly observable, may lack measurement techniques universally used by researchers, or have no available GWAS data.

Examples of latent exposures include known biological mechanisms/concepts, such as chronic inflammation, organ functions, stress, and intelligence level, and unknown underlying processes, for example, some latent factors that are believed to cause different subtypes of psychiatric disorders, functions of gene pathways/modules, and neurological functions in different brain regions. Generally, there are two types of scenarios that often involve latent exposures. First, while data for multiple biomarkers (e.g., inflammation biomarkers like C-reactive protein and cytokines) are available, the actual exposure of interest is some factor that is known to regulate the biomarkers (e.g., chronic inflammation). The other scenario is that, while some observed traits (e.g., metabolites) may pose signals of effects on the outcome, there is evidence that there may be unknown latent features (e.g., lifestyles, diet) underlying the traits that are likely the true risk factors of disease progression. Investigating causal pathways among these latent exposures beyond the level of observable traits may thus yield new findings on causal mechanisms among human traits and diseases.

MR analysis on latent exposures was recently highlighted and investigated(25). To test for the causal effect of a latent exposure on an outcome, one typically relies on GWAS summary statistics for observed traits that are regulated by the exposure. A naive approach is to conduct MR analysis on each single trait and combine evidence of causality across traits. Relying on correlated traits, however, can lead to bias and a loss in power for the analysis due to the effects of horizontal pleiotropy, as the selected IVs may not be truly associated with the latent exposure of interest but only affect the observed traits. For example, the significance and identified direction of causal effect may be inconsistent between different traits. MRLE, the first MR analysis approach designed for latent exposures (25), was recently proposed to improve the validity and power of the analysis by accounting for such pleiotropy between traits. MRLE involves developing a set of estimating functions based on the second-order moments of GWAS summary statistics for the observed traits and the outcome within a structural equation modeling (SEM) framework. In this model, genetic variants are assumed to have indirect effects through latent exposure or possible direct effects on the traits, which allows for the consideration of pleiotropic effects among the traits. Being the first MR analysis approach developed for latent exposures, MRLE has some methodological and computational limitations. First, it is designed for the analysis of a single latent exposure. In many cases, however, we will encounter multiple latent factors behind the observed traits that are potentially correlated, where we will need a flexible approach analogous to multivariable MR that allows us to jointly analyze the direct effects of these latent exposures while accounting for horizontal pleiotropy across these exposures. The SEM framework in MRLE would become overly complex if extended to more complex pleiotropy settings involving multiple correlated latent exposures. Furthermore, we have identified potential convergence issues of the regularized Newton-Raphson’s algorithm for the implementation of MRLE.

In this study, we propose a novel approach for Causal analysis of Latent exposures using Mendelian Randomization (CaLMR). CaLMR adopts a two-sample MR framework and conducts Bayesian modeling based on the likelihood of the estimated SNP-trait associations from summary-level data of the outcome and the observable biomarkers associated with the latent exposures from separate GWAS. CaLMR allows simultaneous testing of the direct causal effects of multiple correlated latent exposures on an outcome, i.e., CaLMR (Multi), with an alternative version, CaLMR (Uni), for analyzing a single latent exposure. In the multiple-exposure scenario, if the exposure-biomarker matchings are known, then we can directly conduct MR analysis based on the known causal diagram. But if we believe there are multiple latent factors that regulate different subgroups of biomarkers, we propose to first conduct a factor analysis using algorithms like genomicSEM(17; 16) to identify the latent factors and learn the underlying exposure-biomarker relationships, based on which we can then apply our MR analysis framework. We show by simulations the validity and efficiency of CaLMR in different data scenarios, where CaLMR (Uni) successfully addresses the convergence issue of the MRLE algorithm in the single-exposure setting, while CaLMR (Multi) effectively identifies the direct causal effects on the outcome among multiple correlated latent exposures and can handle the scenario in which some biomarkers are associated with multiple latent exposures. We also demonstrate the superiority of CaLMR (Uni) to the MRLE algorithm in terms of computational stability, as well as the relative superiority of CaLMR (Multi) to CaLMR (Uni), MRLE, and simple multivariable MR approaches, MVMR-PRESSO and MVMR-IVW, in terms of power and type I error control in the multiple-exposure setting. We further employ CaLMR (Uni) and CaLMR (Multi) to assess the causal effects of four broad psychiatric factors, each indicating shared risk for several related psychiatric disorders, on various autoimmune, digestive, cardiometabolic, or neurodevelopmental diseases. We show that CaLMR can be used to help explain the complexity of the relationship between several psychiatric illnesses regarding their potentially causal effects on numerous other diseases.

## 2 Results

### 2.1 Method Overview

We introduce CaLMR, a Bayesian MR approach to testing the direct causal effect of latent exposures. CaLMR infers the direction and existence of causal effects of unobserved, latent exposures on an outcome leveraging GWAS summary data for the outcome and a series of measurable traits or biomarkers known or hypothesized to be co-regulated by the exposures. Suppose we are interested in analyzing the causal effects of *L* unobserved, latent exposures, *X*_1_, *X*_2_, …, *X*_*L*_, on a disease outcome *Y* utilizing GWAS summary statistics on *M* selected genetic IVs (*G*_1_, *G*_2_, …, *G*_*M*_) for *Y* and *K* observed traits, *B*_1_, *B*_2_, …, *B*_*K*_. The causal diagram in Figure 1(a) describes the assumed relationship between these different variables in the *L* = 2 latent exposure case. The biomarker is deemed valid if (1) it is regulated by at least one of the exposures, and (2) conditional on the exposures, it does not affect the outcome, typically referred to as a “pure surrogate”. We assume the following models for the individual-level data:

**Figure 1:**
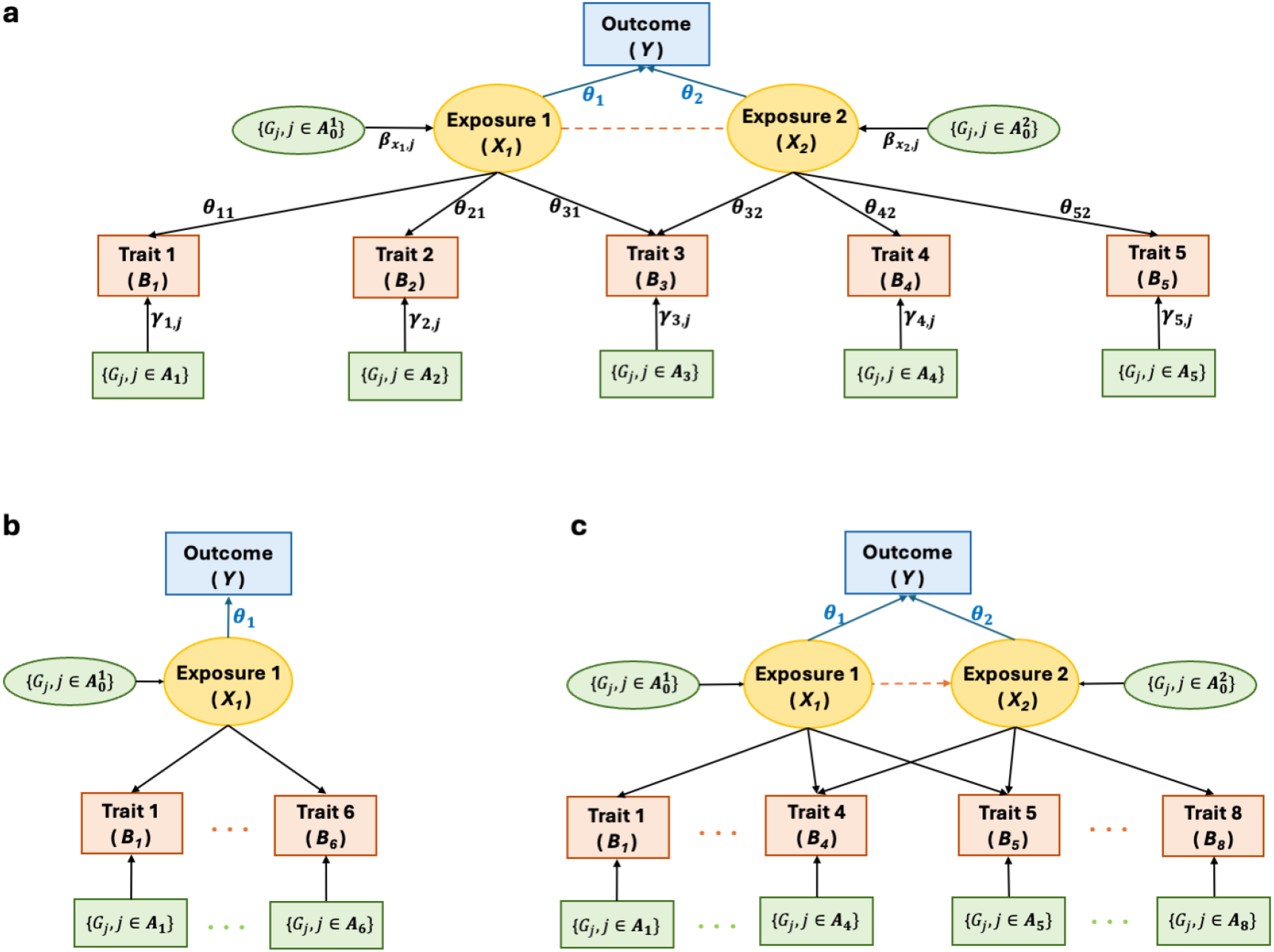
Causal diagrams illustrating the causal links between the outcome (*Y*), latent exposures (*X*_1_ and *X*_2_), observable traits (*B*_*k*_), and genetic IVs (*G*_*j*_). **a**. An example causal diagram describing the general relationship between variables assumed by CaLMR in the two-exposure scenario. *θ*_1_ and *θ*_2_ are the parameters of interests, which are the causal effects of the latent exposures on *Y*. The red dashed line that links the two exposures indicates a potential between-exposure correlation. ***A***_***k***_ denotes the set of SNPs associated with *B*_*k*_ and 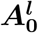 the set of SNPs associated with latent exposure *X*_*l*_. *γ*_*k,j*_s and 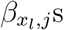 are the direct SNP effects on the observable traits and latent exposures, respectively. The existence of *θ*_31_ and *θ*_32_ reflects the case where some traits (*B*_3_) are regulated by multiple exposures. **b**. A causal diagram describing the single-exposure simulation setting assuming a total of *K* = 6 observable traits. **c**. An illustration of the multiple-exposure simulation setting assuming a total of eight observable traits co-regulated by two correlated latent exposures (correlation: *κ* = ™0.5), among which two (*B*_4_ and *B*_5_) are regulated by both exposures.

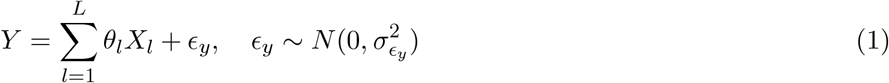

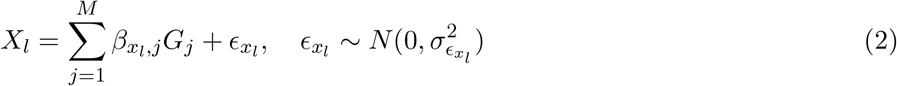

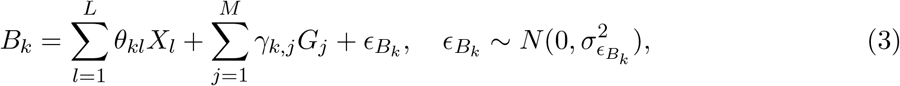

where *θ*_*l*_s are the direct effects of the latent exposures *X*_*l*_s on the outcome, of which we are interested in the significance and signs (positive or negative effects). The rationale of proposing CaLMR (Multi) for jointly analyzing multiple latent exposures is similar to that of the multi-variable MR (MVMR) (5; 44; 43), which is to tackle the issue of horizontal pleiotropy between exposures and increase the testing power. For a set of independent exposures, which, in the MVMR setting, is referred to as “causally independent” exposures(5), implementing CaLMR will be analogous to applying the single-exposure MRLE simultaneously to each of the exposures but will further allow the incorporation of pleiotropic IVs, i.e., IVs that have horizontal pleiotropy between exposures, which could yield better inference. In the case of correlated (i.e., “causally dependent”) latent exposures (Figure 1(a), with the red dashed line), however, univariate MR analysis of each exposure will examine not the direct effect of each exposure on the outcome but “total” causal effect 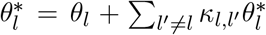 the sum of its direct effect and the indirect effect through its association with other causal exposures, where 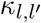 denotes the correlation between exposures *l* and *l*^′^, 1 ≤ *l, l*^′^ ≤ *L*. This may lead to inflated false discoveries on noncausal exposures due to correlations with other exposures. Like MVMR methods, CaLMR (Multi) is designed to provide unbiased estimates of the direct causal effects of the latent exposures *θ*_*l*_s by accounting for between-exposure correlations. Note that the CaLMR (Multi) method will reduce to CaLMR (Uni) in the case of *L* = 1.

We assume that the selected genetic IVs (*G*_*j*_s) may have direct effects on both the exposures 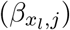 and the observed traits (*γ*_*k,j*_). We assume independence between 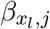 and *γ*_*k,j*_, i.e., uncorrelated pleiotropy, in the prior distributions for these parameters. But it has previously been shown that the modeling framework is robust to correlated pleiotropy between these traits. We propose a strict criterion for the selection of IVs used in the analysis. Adopting the IV selection procedures established for MRLE and MVMR, we select SNPs that are significantly associated with at least two constituent biomarkers for each exposure and then take the union of these SNP sets across all exposures. This is to ensure a high chance of selecting the SNPs that are truly associated with the exposures. Our current model assumes no direct effect of IVs on *Y*, i.e., no horizontal pleiotropy between the exposures and the outcome. Such horizontal pleiotropy can be easily accounted for by directly incorporating a recently developed approach into our Bayesian modeling framework (9). The stringent IV selection criterion with a genome-wide significance threshold (p=5e-8) may lead to insufficient or, sometimes, no IVs selected for the analysis. To ensure a sufficient amount of IVs are included in the analysis, we propose a data-driven approach for IV selection. Specifically, we initially set the significance threshold to *p*_0_ = 5 ×10^−8^. The selection process iterates, with the significance threshold doubled at each step until the number of IVs selected for each exposure reaches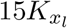, where 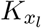 denotes the number of observable traits associated with exposure *l*.

To infer the direction of the causal effects *θ*_*l*_s, CaLMR requires that the signs of *θ*_*k,l*_s (the exposure-biomarker associations) are known. This is a reasonable assumption: for the well-defined latent exposures, the directions of the effect of exposures (e.g., kidney function) on the observed traits (kidney function biomarkers) are well-known or documented in the literature, while for the unknown latent exposure, algorithms like genomicSEM can provide an estimate of such directions. We want to note that if the signs of exposure-trait associations are unknown, then CaLMR can still provide a hypothesis testing result on the existence of causal effects but without inference on their directions. CaLMR conducts inference on the parameters based on a Markov chain Monte Carlo (MCMC) algorithm with Gibbs sampling. Details of the Bayesian inference and the MCMC algorithms for CaLMR (Uni) and CaLMR (Multi) are provided in the Methods section. Derivation of the MCMC algorithms and discussion on identifiability of the inferred signs of the causla effects and between-exposure correlations are provided in Section 1 of the Supplementary Materials.

### 2.2 Simulation Studies

In the single-exposure setting, we showed by simulation that CaLMR (Uni) and MRLE had comparable powers in detecting the causal effect of the exposure, which is expected because the two approaches utilize the same data information. Using *p* = 5*e* − 4 and 5*e* − 5 as the significance threshold for IV selection leads to well-controlled type I error rates at similar levels around *α* = 0.05 for CaLMR (Uni), suggesting that CaLMR (Uni) is robust to the significance threshold for IV selection (Figure 2a, Supplementary Table 1). When GWAS sample size is relatively small, CaLMR (Uni) tends to have a higher power when using a less stringent (but reasonable) significance threshold for IV selection to include more IVs in the analysis. We also compared CaLMR with two standard MR methods for observable exposures, MR-PRESSO(52) and MR-IVW. Since MR-PRESSO and MR-IVW are not designed for analysis on latent exposures, we apply them to conduct a test on each observed trait with an adjusted p-value significance threshold at 1− (1 − 0.05)^1*/K*^ and concluded that the latent exposure had a significant effect on the outcome if any of the tests had a significant result. Results suggested that MR-PRESSO and MR-IVW lacked sufficient power to identify the causal latent exposure, which is consistent with the findings reported in the paper for MRLE (25).

**Figure 2:**
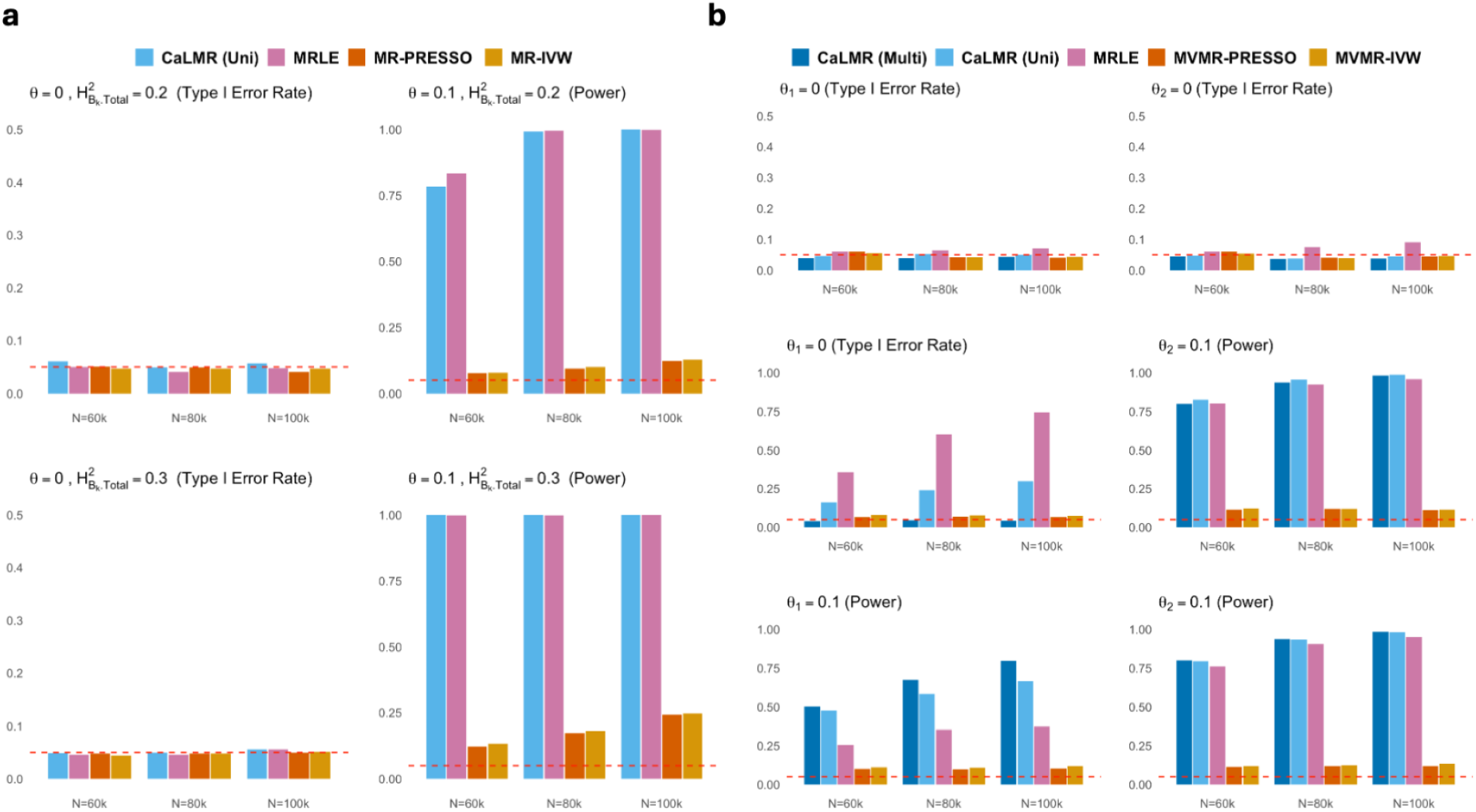
Simulation results under the single-exposure (**a**) and multiple-exposure (**b**) scenarios assuming a GWAS sample size for each trait and the outcome *N* = 60K, 80K, or 100K. Results are summarized across 1000 simulations per setting. **a**. type I error control and power comparison between CaLMR (Uni), MRLE, MR-PRESSO, and MR-IVW under the single exposure setting. The significance threshold for IV selection is equal to *p* = 5 ×10^−4^. We assume the heritability of *X* is 0.2, and the total heritability of each biomarker 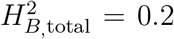 (top panel) or 0.3 (bottom panel). **b**. type I error control and power comparison between CaLMR (Multi), CaLMR (Uni), MRLE, MV MR-PRESSO, and MVMR-IVW in the setting of two correlated latent exposures.

Our simulation results also showed the superiority and robustness of CaLMR (Multi) compared to alternative methods in the multiple-exposure settings. We assumed there were two latent factors *X*_1_ and *X*_2_ with a correlation -0.5. We generated summary statistics for *K* biomarkers and the outcome based on models (1-3) assumed by CaLMR (details are summarized in the Methods section). We applied the data-driven approach described in Section 2.1 to select IVs. We compared CaLMR (Multi) with four alternative methods, including CaLMR (Uni), MRLE, MVMR-PRESSO, and MVMR-IVW. The latter two methods are the multivariable versions of MR-PRESSO and MR-IVW implemented in our simulations for the single-exposure setting which were implemented by an ad hoc approach (see Methods). When implementing CaLMR (Uni) and MRLE, we applied the test to each exposure using SNPs associated with that exposure and tested the effects of each exposure separately. We used the union set of IVs selected for the two exposures for the implementation of CaLMR (Multi), MVMR-PRESSO, and MVMR-IVW. To control the family-wise error rate (FWER) at 0.05 for each exposure, when implementing MVMR-PRESSO and MVMR-IVW, we used the adjusted p-value at 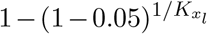 given the multiple hypotheses tests performed. We examined the performance of the various methods in three scenarios: (1) no causal effect from either biomarker, i.e., *θ*_1_ = *θ*_2_ = 0, (2) exposure one was causal and exposure two was not, with *θ*_1_ = 0 and *θ*_2_ = 0.1, and (3) both exposures were causal, with *θ*_1_ = *θ*_2_ = 0.1.

We assumed there were a total of *K* = 8 biomarkers and 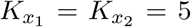,implying two of them were regulated by two exposures simultaneously. CaLMR (Multi), MVMR-PRESSO, and MVMR-IVW controlled the type I error rate sharply (Figure 2b). However, MVMR-PRESSO and MVMR-IVW had extremely low power compared to CaLMR (Multi) for detecting causal exposures in all three scenarios. As the GWAS sample size increased, their power went slightly upward, but they were still overly conservative in identifying causality from the exposures. When there was no causal exposure, CaLMR (Uni) could control the type I error rate well, while MRLE sometimes had a slightly inflated type I error rate, which was due to some convergence issues we previously discussed (See Supplementary Materials). However, when only one of the two exposures was causal, CaLMR (Uni) and MRLE could have a largely inflated type I error rate for the non-causal exposure in the presence of a causal exposure. MRLE tended to have higher inflation in type I error rate than CaLMR (Uni). The inflation persisted as the GWAS sample sizes increased. CaLMR (Uni) and MRLE also had lower power than CaLMR (Multi) when both exposures were associated with the outcome. This is because the negative correlation between the exposures undermined the power of detecting true causal effects when analyzing them marginally. We have shown previously that MRLE is robust to correlated pleiotropy between the observable traits, which suggests that CaLMR, having the same model assumptions, should have similar performance as MRLE in these pleiotropy settings. We have also conducted simulations assuming alternative data scenarios and have summarized the results in Section 2 of the Supplementary Materials.

### 2.3 Analyzing the Causal Effect of Psychiatric Factors on Disease Risks

Severe mental illness has been linked with higher risks of a wide range of chronic physical conditions (47). Studies have shown evidence of causal links between different types of psychiatric disorders on different types of diseases. For example, epidemiological studies have reported the association of a range of psychiatric disorders, such as schizophrenia and psychosis, with genetic markers of the immune system, various immune alterations, and autoimmune diseases, such as systemic lupus erythematosis and multiple sclerosis (MS), in different organs (23; 2). Additionally, some recent studies have revealed shared mechanisms across a series of major psychiatric and neurodegenerative diseases, providing insights into alternative early treatment and therapeutic development for late-life neurodegenerative diseases (13; 53; 42). The possible causal link between psychological disorders and digestive disorders have been identified in various studies, which may be explained by functional abnormalities in the brain-gut axis and the increasingly recognized impact of psychological distress on the systemic and gut immunity(54; 14; 14; 31). Studies have also consistently shown an increased risk of cardiovascular and cardiometabolic diseases associated with depression, schizophrenia, and other severe mental illnesses (20; 1; 38).

Given this evidence and recent research interest in the potential causal effects of psychiatric illnesses (and of disorders afflicting the brain more generally) on various other health outcomes, we have conducted a systematic investigation on the causal signals involving eleven psychiatric disorders on eighteen neurodegenerative, cardiometabolic, digestive, and autoimmune disease outcomes. these relationships systematically (36). We have collected GWAS summary statistics for problematic alcohol use (ALCH), post-traumatic stress disorder (PTSD), anorexia nervosa (AN), and obsessive-compulsive disorder (OCD) from FinnGen(28) and attention-deficit/hyperactivity disorder (ADHD), autism spectrum disorder (AUT), anxiety (ANX), bipolar (BIP), major depression disorder(MDD), obsessive-compulsive disorder (OCD), schizophrenia (SCZ), and Tourette syndrome (TS) from the Psychiatric Genomics Consortium (PGC)(49), totaling *K* = 11 major psychiatric disorders. Figure 3(a) shows the genetic correlation among these psychiatric disorders estimated by LD score regression (LDSC) (4).

**Figure 3:**
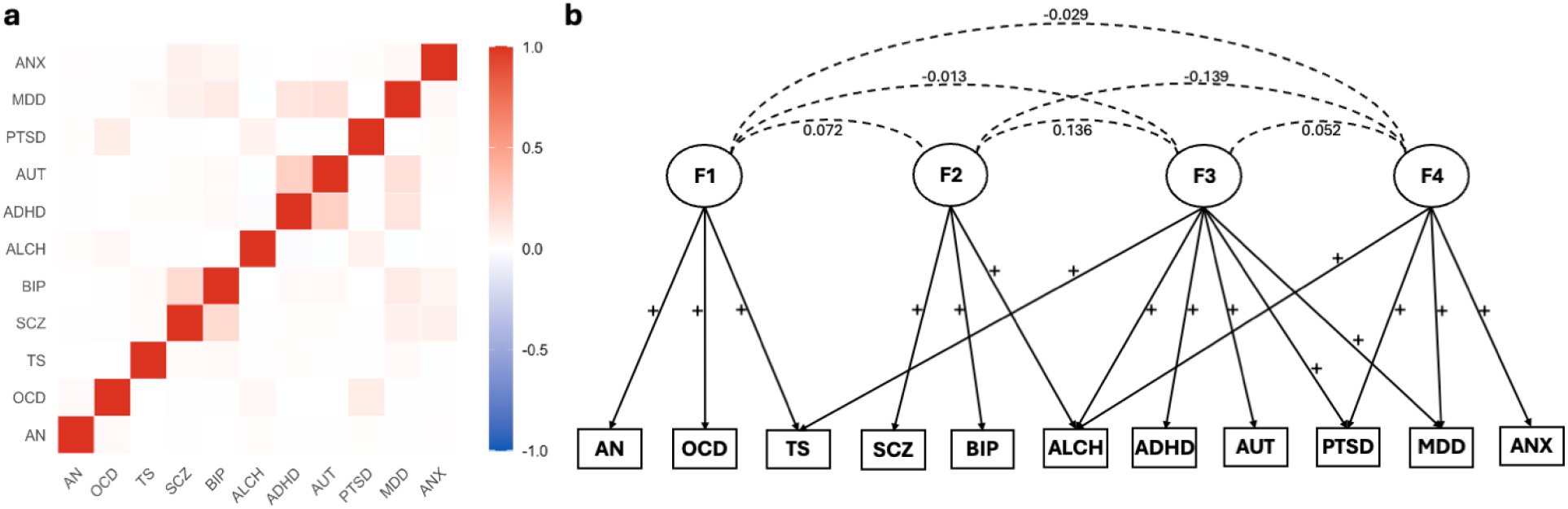
The genetic architecture and identified latent factor structures among eleven psychiatric disorders. **a**. Genetic correlation structure of the eleven disorders estimated using LDSC. **b**. Four latent psychiatric factors underlying the 11 disorders and their associations with the disorders inferred by genomicSEM. The four factors align well with four subtypes of psychiatric disorders, including compulsive, psychotic, neurodevelopmental, and internalizing disorders. The values on the dashed lines connecting each pair of factors are the between-factor correlations estimated by CaLMR (Multi). The signs near the solid lines linking factors with the observed disorders indicate the estimated signs of genetic correlations derived from genomicSEM. The signs reflect our known directions of the exposure-trait associations and will be used to determine the direction of the exposure-outcome effects. Abbreviations: AN (anorexia nervosa), OCD (obsessive-compulsive disorder), TS (Tourette syndrome), SCZ (schizophrenia), BIP (bipolar disorder), ALCH (problematic alcohol use), ADHD (attention-deficit/hyperactivity disorder), AUT (autism spectrum disorder), PTSD (post-traumatic stress disorder), MDD (major depressive disorder), ANX (anxiety).

**Figure 4:**
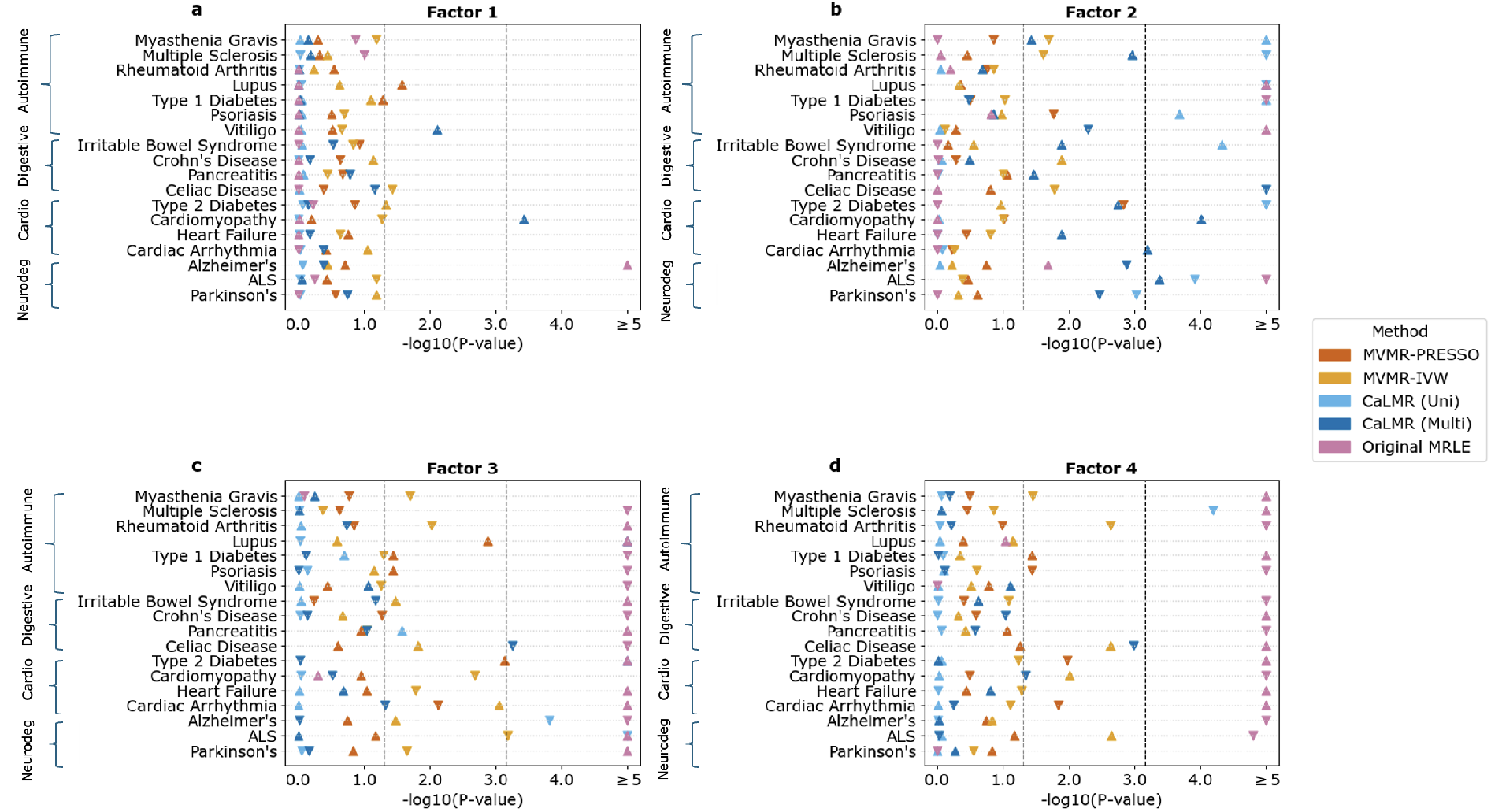
Analysis of Causal Effects of Psychiatric Factors on Various Disease Outcomes. We report the ™ log_10_ p-values associated with testing the causal effects of four latent psychiatric factors on 18 disease outcomes using five methods, including MVMR-IVW, MVMR-PRESSO, MRLE, CaLMR (Uni), and CaLMR (Multi). Upwardpointing triangles represent positive causal estimates, whereas downward-pointing triangles represent negative estimates. The gray dashed line marks *α* = 0.05, and the black dashed line marks the adjusted significance level Z^′^ = 0.0007. Results are summarized by factor for Factor 1 (aligned with compulsive disorders, **a**), Factor 2 (aligned with psychotic disorders, **b**), Factor 3 (aligned with neurodevelopmental disorders, **c**, and Factor 4 (aligned with internalizing disorders, **d**. The eighteen disease outcomes include various autoimmune, digestive, cardiometabolic, and neurodegenerative disorders as indicated by the brackets on the y-axis.

We derived four broad latent psychiatric factors from the eleven disorders using genomic Structral Equation Modeling (genomicSEM)(17) that explain both shared and unique genetic risks across these disorders (18) (Figure 3b). These four factors align well with the four subtypes of psychiatric disorders: compulsive, psychotic, neurodevelopmental, and internalizing disorders (Figure 3b). We thus believe they influence various biobehavioral traits and brain functions and may offer insights into the underpinnings of their associated psychiatric disorders (see Section 3 of the Supplementary Materials).

Following our modeling framework, we treated the psychiatric factors as latent exposures and the set of specific psychiatric disorders associated with each factor as their “biomarkers”. In total, there are *K* = 11 biomarkers across *L* = 4 latent factors, with some biomarkers being associated with more than one latent factor. We applied both the CaLMR (Uni) and CaLMR (Multi) methods, along with MRLE, MVMR-PRESSO, and MVMR-IVW. When applying MRLE and CaLMR (Uni) in a single-exposure setting, a different IV selection threshold was used for each psychiatric factor. This was done to ensure that there were a sufficient number of IVs used for each factor. Specifically, p-value thresholds of 5 ×10^−3^, 5 ×10^−6^, 5 ×10^−4^, and 5 ×10^−3^ were employed for factors 1, 2, 3, and 4, respectively, that correspond to compulsive, psychiotic, neurodevelopmental, and internalizing disorders. For CaLMR (Multi), MVMR-PRESSO, and MVMR-IVW, IVs were selected at a liberal p-value threshold of 5 ×10^−3^ across all four factors.

This was done to ensure all four factors had a sufficient number of significant IVs selected for the joint analysis so that the results would not be biased towards any of the factors. We selected independent IVs by performing linkage disequilibrium (LD) clumping on the remaining SNPs using a 0.5 MB window and an *r*^2^ threshold of 0.005, implemented in PLINK. For each method, we tested the null hypothesis of no significant causal effect from the latent factor on the outcome at the *α* = 0.05 significance level. We also compared the p-values to the adjusted significance threshold of *α*^*t*^ = 1 − (1 − 0.05)^1*/*72^ ≈ 0.0007 as there were in total 72 tests conducted for each method.

Figure 3 illustrates the resulting p-values from our analyses. In all scenarios, CaLMR (Uni) and CaLMR (Multi) performed the best at identifying potential causal effects. We found that the MRLE method had a much higher significance than other methods, particularly when testing the effects of factors 3 and 4 (16 and 15 out of the 18 tests produced *p <* 10^−5^, respectively), which may indicate highly inflated false positive rates. Comparatively, CaLMR (Uni) produced significant results for two outcomes related to factor 3, while the CaLMR (Multi) rejected the null for one outcome when testing factor 3 after adjusting for the effects of the other factors. Additionally, CaLMR (Uni) showed a significant result for only one outcome when testing the direct causal effect of factor 4, whereas the CaLMR (Multi) detected no causal effect of factor 4 on any of the outcomes. In comparison, the MVMR-PRESSO and MVMR-IVW both appear to be conservative, which is consistent with our simulation results where they showed low power. Between both methods and for all four factors, the only significant causal effect detected was from factor 3 on ALS using MVMR-IVW, which could be a false discovery given the high false positive rates typically associated with the IVW test. This agrees with the results of our simulation studies, where both MVMR-PRESSO and MVMR-IVW appeared to have exceedingly low power while MRLE suffered from an inflated type I error.

The superior control of type I error by the CaLMR (Uni) method, compared to the MRLE method when testing for causal effects from factors 3 and 4, may be attributed to the fact that these two factors are tested with *K >* 3 biomarkers. Specifically, factor 3 has *K*_3_ = 6 biomarkers, while factor 4 has *K*_4_ = 4 biomarkers. For these two factors, the MRLE method required substantially more time to complete the analysis and frequently issued warning messages related to the singularity of some asymptotic covariance matrix. In contrast, CaLMR (Uni) took only marginally more time to analyze factors 3 and 4 compared to factors 1 and 2, and did not exhibit any performance issues during these analyses.

In addition to the testing on factors 3 and 4, CaLMR (Uni) detected causal effects from factor 2 on 8 out of 18 outcomes, while CaLMR (Multi) detected causal effects on 4 outcomes from factor 2. This difference in causal detections likely stems from the use of relatively weak instruments by CaLMR (Multi) given the less stringent p-value threshold used for IV selection. Another potential reason for the apparently lower significance of CaLMR (Multi) relative to CaLMR (Uni) is that it uses the union set of all IVs valid for each individual latent factor, of which only a small portion are valid for factor 2; this pattern was also observed in our simulations. Factor 2 indicates general psychotic disorder and has *K* = 3 associated biomarkers which are Schizophrenia (SCZ), Bipolar Disorder (BIP), and ALCH. The other latent factors studied describe compulsive disorders (factor 1), neurodevelopmental disorders (factor 3), and internalizing disorders (factor 4). An individual prone to psychotic disorder is especially concerning compared to other types of disorders because they may lose touch with reality entirely. This can severely impact their social life, career, and ability to manage their health, among other aspects, leading to further destabilization in their life (30). It should also be noted that antipsychotic drugs can be particularly toxic and disrupt many bodily functions (48). Thus, researchers might reasonably anticipate that psychotic disorders, by their nature, increase the likelihood of developing other disorders in affected individuals.

Among the eight outcomes determined to be causally affected by factor 2, five were classified as autoimmune disorders, while the remaining three belonged to each one of the other outcome categories. We caution that there may have been issues concerning identifiability as among the significant causal estimates, those estimates for Irritable Bowel Syndrome (IBS), Type 1 Diabetes (T1D), Psoriasis, and Myasthenia gravis were positive while estimates for ALS, Multiple Sclerosis (MS), Type 2 Diabetes (T2D), and Lupus were negative. It is plausible that the causal effect on ALS from factor 2 would be negative since ALS is believed to be partially characterized by a dopaminergic deficit while psychotic illness is characterized by a dopaminergic excess (51) (22). However, negative causal effects on MS, T2D, and Lupus contradict the existing literature concerning the relative comorbidity of SCZ with these diseases (50) (55).

These significant results are consistent with the existing literature on the relationship between psychotic and autoimmune disorders. Previous research has suggested that a family history of psychotic disorders is linked to a higher risk of developing autoimmune diseases, and the reverse also appears to be true (23). Additionally, inflammatory markers have been found at elevated levels in the blood and cerebrospinal fluid (CSF) of individuals with psychosis, with particularly high concentrations in cases of first-episode psychosis. Furthermore, a recent meta-analysis has suggested that the risk of developing an autoimmune disease was 55% greater in individuals with a previous diagnosis of a psychotic disorder (11). The results produced from our CaLMR (Uni) method should help to clarify the nature of the relationship between psychotic and autoimmune diseases.

## 3 Discussion

We introduce CaLMR as a new method that jointly tests for the causal effects of latent exposures on an outcome. Compared to MRLE, the single-exposure method for latent exposure, the single-exposure version of our method, CaLMR (Uni), can accurately detect causal effects at a similar level of power while offering faster computation time and controlling type I error much better, particularly in cases where the number of traits associated with the exposure is more than three. Our results show that strictly selecting IVs associated with at least two observable traits corresponding to each latent exposure is sufficient in determining causal effects. When testing for effects of multiple latent exposures simultaneously, existing ad-hoc approaches, i.e., combining signals from trait-specific MR analyses, had very low power due to the inefficient way of information integration and reliance on the union of the set of valid IVs for each exposure tested. We showed that CaLMR (Multi) did not suffer nearly as much of a reduction in power when compared to these methods. Additionally, CaLMR (Multi) is robust in scenarios in which there are many latent exposures and relatively few IVs in comparison.

We studied the causal effects of four latent psychiatric factors on various disease outcomes using CaLMR and alternative MR methods. CaLMR (Uni) detected causal effects from the latent psychotic factor on five out of seven autoimmune disorder outcomes. While these significant causal effects corroborate recent research, we stress the need to consider possible confounding in these results. To adjust for residual confounding, we excluded SNPs associated with potential confounders—including sleep disorders, obesity, and pro-inflammatory cytokines—based on

GWASs obtained from the UK Biobank, applying a significance threshold of 5 ×10^−6^. Too few SNPs were removed to make a discernible difference in the results and we further acknowledge the inability to adjust for certain confounders like medication use.

One thing to note is that correct identification of the direction of the causal effects by CaLMR requires correct specification of all biomarker-exposure association directions in the analysis. While the directions of these associations are frequently known and can be easily obtained from publicly available literature, issues may arise as the number of latent exposures and associated biomarkers grows. Nevertheless, CalMR still provides more reliable conclusions on the direction of the effects than traditional approaches that may not be able to draw solid conclusions on the direction of the effect with opposite directions of the trait-outcome relationship detected on different traits, and CaLMR (Multi) further refine the analysis by reducing potential bias due to horizontal pleiotropy and correlation among the latent exposures. This could also explain the smaller number of discoveries by CaLMR (Multi) than CaLMR (Uni) in our results. As with any MR method, low p-values of the CaLMR tests are not to be regarded as necessarily proving the existence of causal effect(s) from the latent exposure(s) on the outcome but rather as suggesting a possible causal signal that warrants further investigation. Such results could be influenced by confounding, other types of pleiotropy that are not accounted for, or instrument strength, and therefore, further attention must be directed toward assessing and mitigating these influences before drawing solid conclusions about causal relationships. Some extensions of CaLMR can be considered to further improve the testing power. We currently only incorporate independent IVs as in a standard MR framework, but CaLMR can further incorporate correlated SNPs as IVs by accounting for LD among significant SNPs in the likelihood for GWAS summary statistics (9). This could reduce weak instrument bias and improve power by incorporating more significant GWAS signals with a more stringent significance threshold for IV selection. This extension can be quite useful, especially when the GWAS sample sizes for exposures are relatively small and the GWAS signals are relatively weak. CaLMR currently does not account for horizontal pleiotropy between the latent exposures and outcome, but such pleiotropy can be accounted for by directly incorporating a recent approach to dealing with correlated pleiotropy (9) into our modeling framework.

Looking ahead to future research, we stress the growing interest in causal inference within the context of integrative multi-omics analysis, which has garnered increasing attention, especially in recent years(46; 35; 21; 7; 27). The growing availability of public multi-omic data sources on genomics, metabolomics, proteomics, and other molecular datasets poses valuable opportunities for us to uncover causal biological processes underlying disease progression and discover new drug targets. While a few MR methods have been proposed for jointly analyzing the causal effects of proteins and metabolomics(8; 7), MR integrating GWAS information on high-dimensional exposures is still an understudied but critical topic. The extension of our CaLMR framework to the high-dimensional-biomarker setting could contribute to the field by revealing potential lower-dimensional latent causal pathways beyond the level of observed, high-dimensional biomarkers.

## Data Availability

All data produced in the present study are available upon reasonable request to the authors.

## 4 Acknowledgements

This work was supported by the National Institutes of Health (NIH) grant R00 HG012223 (J.J.).

## Methods

### Model setup

#### CaLMR (Uni) for the Single-Exposure Case

In the one latent exposure setting, Models (1 - 3) can be simplified to:

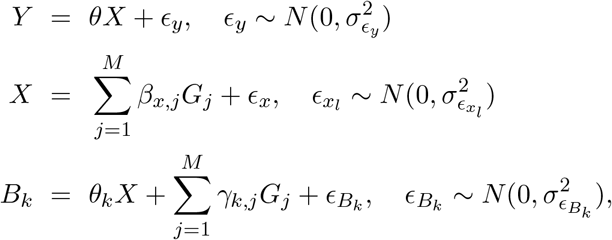

where we assume a direct effect of genetic IV *G*_*j*_ on *X* with a prior 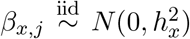,with 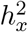 denoting the per-SNP heritability of *X*. Each observable trait *B*_*k*_ is assumed to be explained by both *X* and *G*_*j*_s, with the direct effect from *G*_*j*_ following a prior distribution 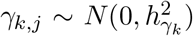, *j* = 1, …, *M, k* = 1, …, *K*.

CaLMR (Uni) conducts inference under a Bayesian modeling framework using an MCMC algorithm with Gibbs sampling on model parameters, 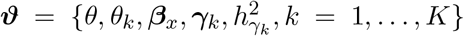.

We use conjugate priors for parameters, which enable us to derive the closed form of the full conditional distributions of the parameters and conduct Gibbs sampling with slight adjustments on some of the steps to approximate the joint posterior distribution. Throughout the text, we conduct inference on the standardized scale, i.e., *G*_*j*_s, *X, B*_*k*_s, and *Y* are standardized to have a zero mean and unit variance. We develop CaLMR (Uni) with the following assumptions: (a) independent IVs, (b) no horizontal pleiotropy between the exposure and the outcome, and (c) small *K* setting. The Regression with Summary Statistics (RSS) Likelihood of the *M* genetic 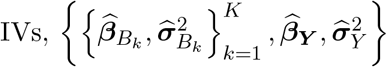, is:

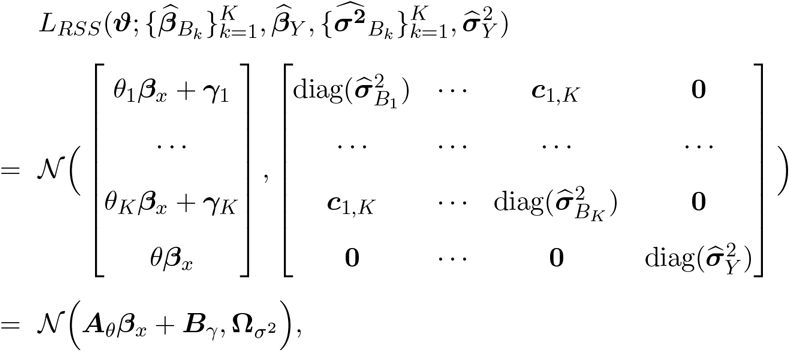

where 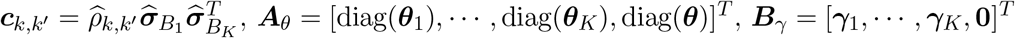,and 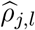 denotes the estimated genetic correlation between biomarker *B*_*j*_ and *B*_*l*_ from the intercept in bivariate LD score regressions. We assume the following conjugate priors for the parameters: 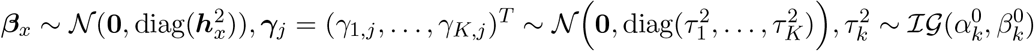,where 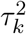 is the prior variance of 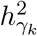, and a non-informative prior for ***η***_*θ*_ = (*θ*_1_, …, *θ*_*K*_, *θ*)^*T*^.

#### CaLMR (Multi) for the Multiple-Exposure Case

In practice, there may be multiple latent factors underlying the observed traits, and CaLMR (Uni) can be extended to the multiple-exposure settings, which we refer to as CaLMR (Multi), to conduct joint analysis accounting for potential horizontal pleiotropy and correlation between the exposures. First, for the latent exposure *X*_*l*_, *l* = 1, …, *L*, assume it regulates *K*_*l*_ different biomarkers. Like the assumptions we made in CaLMR (Uni), we suggest the SNPs *G*_*j*_s, the biomarkers *B*_*k*_s, the latent exposures *X*_*l*_s, and the outcome *Y* are all standardized. Recall the three proposed models are

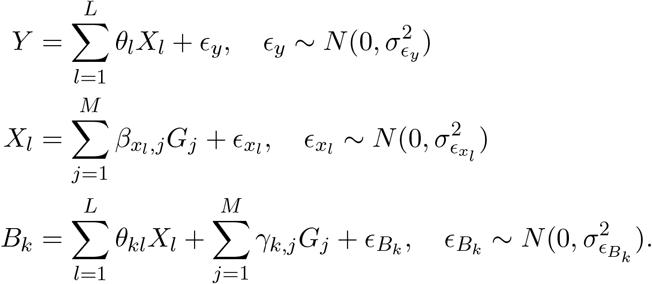

Without loss of generality, 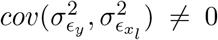 and 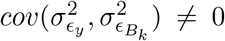 due to the existence of potential confounders. The prior for the direct effect of genetic IV *G*_*j*_ on *X*_*l*_ is 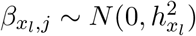, with 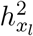 represents the per-SNP heritability of exposure *X*_*l*_. The genetic variability of each observable trait *B*_*k*_ is assumed to be explained by its associated latent exposures and genetic variants *G*_*j*_s. We assume the direct effect from *G*_*j*_ has prior 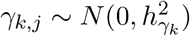.

CaLMR (Multi) is also implemented under the Bayesian framework using an MCMC algorithm to generate posterior samples of the model parameters 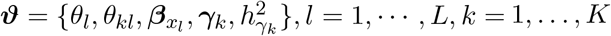. The full conditional distributions of these parameters are derived using conjugate priors, and we generate posterior samples using Gibbs sampling approach. Under the same assumptions as CaLMR (Uni), we derive the Regression with Summary Statistics (RSS) Likelihood of the *M* genetic variants, 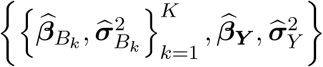,as

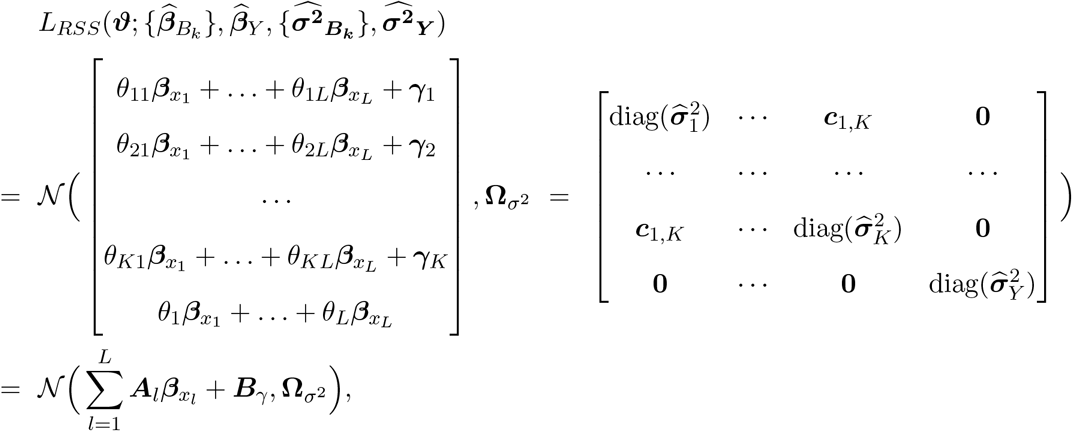

Where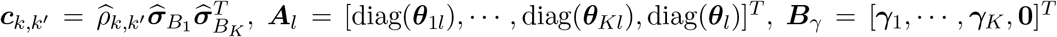, and 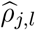 denotes the estimated genetic correlation between biomarker *B*_*j*_ and *B*_*l*_ from the intercept in bivariate LD score regressions. We assume the conjugate priors for the parameter: 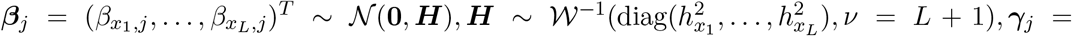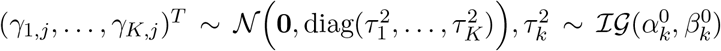, where 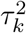 is the prior variance of 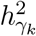, and non-informative priors for *θ*_*kl*_ and *θ*_*l*_s.

### Generating GWAS Summary Statistics in the Simulation Studies

#### Single-Exposure Setting

In the simulation studies, we generated the GWAS summary-level statistics directly to work on. For all our analyses, we used the 1000 Genomes Phase 3 European Sample as the reference data to conduct LD clumping to select independent IVs and used the 1.2 million SNPs in HapMap3 SNP list to conduct the tests. Suppose there are *M*_0_ = 2 ×10^5^ total independent SNPs representing common variants throughout the genome. We set equal GWAS sample sizes for the outcome and K observed traits. For any pair of biomarkers, *B*_*k*_ and *B*_*l*_, *k /*= *l*, we assume the overlapping sample size is 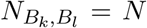 and the genetic correlation between the two is *ρ*_*l,m*_ = *cor*(*B*_*l*_, *B*_*m*_) = *±*0.3.

Importantly, we assume no overlapping samples between any biomarker *B*_*k*_ and the outcome *Y* to avoid the pleiotropy situations, i.e., 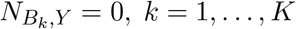.We further assumed that 1% of the SNPs are truly associated with latent exposure *X* and each trait *B*_*k*_. The total heritability of each trait is established at 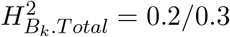, where 20% of them is explained by the latent exposure 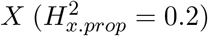. The effects of *X* on each observed trait is set equal at 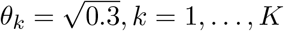.Therefore, we derive the following equations and distributions for SNP j:

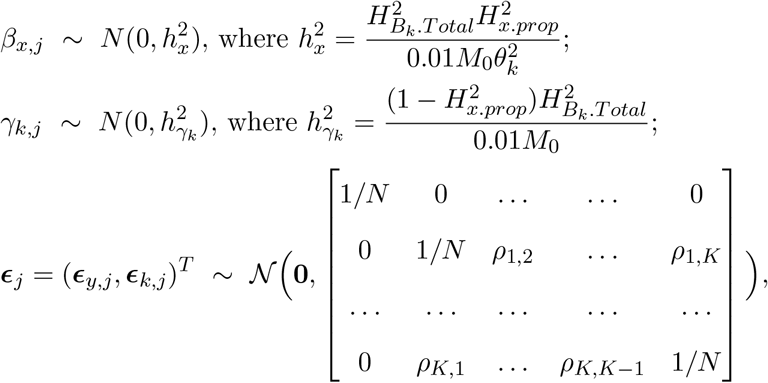

where *k* = 1, …, *K*. The summary coefficients for *B*_*k*_ and *Y* are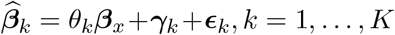, and 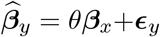;and the standard errors for *B*_*k*_ and *Y* are 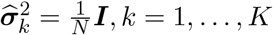 and 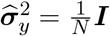.

#### Multiple-Exposure Setting

In the multiple-exposure simulation, we assume there exist two correlated latent exposures underlying *K* observed traits. We further consider the GWAS for *B*_*k*_s have no overlapping samples so that *ρ*_*l,m*_ = *cor*(*B*_*l*_, *B*_*m*_) = 0, *l /*= *m*. Similar to the single-exposure settings, we assume 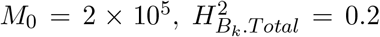, and 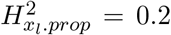 for each *X*_*l*_, and each latent factor *X*_*l*_ explains 30% of the variability of each biomarker it regulates, i.e., 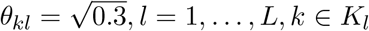.Additionally, we assume there exists an initial causal pathway between *X*_1_ and *X*_2_, with 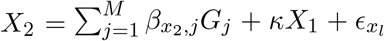, where *κ* denotes the correlation between the two exposures (Figure 1c). Given this causal pathway, the total effect of *G*_*j*_ on the outcome *Y* that go through *X*_2_ is 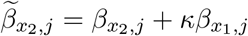.For each SNP, we have

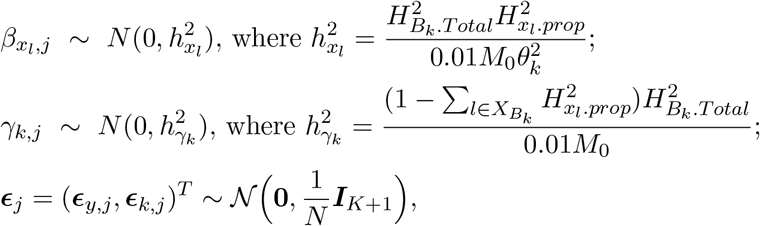

where *j* = 1, · · ·, *N* and *k* = 1, …, *K*. The summary coefficients for *B*_*k*_ and *Y* are 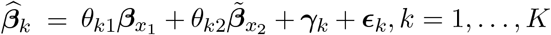 and 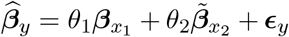;the standard errors for *B*_*k*_ and *Y* are 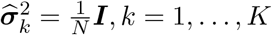 and 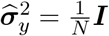.

#### Analyzing the Causal Effects of Psychiatric Factors on Disease Risks

We obtained GWAS summary statistics for each of the eleven psychiatric disorders primarily from the Psychiatric Genomics Consortium (PGC), which is the largest psychiatric consortium in existence. To ensure that the strict condition of no overlap between the biomarkers and outcomes is satisfied, for some of the biomarkers, we rely on summary data obtained by FinnGen, which consists of data obtained from 500,000 Finnish biobank participants. This was done for biomarkers whose PGC samples appear to have some overlap with UK Biobank, which is unacceptable since GWAS summary data for all 18 disease outcomes were obtained from UK Biobank. All datasets were converted to Genome Build 37 unless they were already in that format. Additional preprocessing steps were applied to each dataset including renaming columns to ensure common names, removing SNPs with minimum allele frequency (MAF) below 0.01, removing multiallelic SNPs, converting odds ratio estimates into covariate estimates, realigning all SNPs to have the same allele order across all datasets, and calculating effective sample size if not already available.

Given that summary-level data for the 11 psychiatric disorders had overlapping samples, as all of them are sourced either from PGC or FinnGen, we estimated the between-biomarker covariance by applying bivariate LDSC before conducting our analyses (Figure 3a). Furthermore, while the original MRLE method provides p-values for testing the null hypothesis of no causal effect along with the direction of *θ*, CaLMR (Uni) and CaLMR (Multi) produce a 95% credible interval for *θ*. To calculate the Bayesian p-value based on posterior samples, we use the following expression:

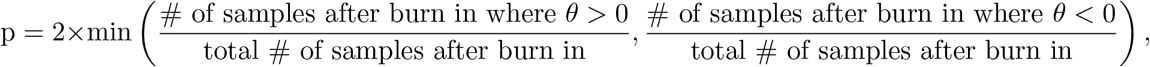

which estimates the probability 2 ×min*{P* (*θ >* 0), *P* (*θ <* 0)*}*.

## 5 Ethics declarations

### Competing interests

The authors declare no competing interests.

## 6 Data Availability

The 1000 Genomes Phase 3 genotype data used in our analysis can be accessed at https://data.broadinstitute.org/alkesgroup/LDSCORE/. All GWAS summary statistics used in this study are publicly available. GWAS summary statistics for ALCH, PTSD, AN, and OCD can be downloaded from the FinnGen website at https://www.finngen.fi/en. GWAS summary statistics for MDD, ADHD, BIP, ANX, AUT, and TS can be downloaded from the PGC website at https://pgc.unc.edu/for-researchers/download-results/. GWAS summary statistics for the eighteen disease outcomes and confounders were obtained from the UK Biobank and can be downloaded from the GWAS Catalogue website at https://www.ebi.ac.uk/gwas/.

## 7 Code Availability

The R package “CaLMR” is available at https://github.com/yueuuy/CaLMR/.

## Notes

### Competing Interest Statement

The authors have declared no competing interest.

