## Supplementary Materials for "Bayesian Mendelian Randomization Analysis for Latent Exposures Leveraging GWAS Summary Statistics for Traits Co-Regulated by the Exposures"

#### 1 MCMC algorithm for CaLMR

##### 1.1 CaLMR (Uni) for Single Latent Exposure Analysis

In order to have the full conditional distributions for the parameters of interest, we first derive the "Regression with Summary Statistics" (RSS) likelihood for the summary data. In the single-exposure setting, recall the models are simplified to:

$$
\begin{aligned}
Y &= \theta X + \epsilon_y, \\
X &= \sum_{j=1}^{M} \beta_{x,j} G_j + \epsilon_x, \\
B_k &= \theta_k X + \sum_{j=1}^{M} \gamma_{k,j} G_j + \epsilon_{B_k}, k = 1, \ldots, K,
\end{aligned}
$$

where $Y$, $X$, and $B_k$ denotes the outcome, latent exposure, and observable traits, respectively. Regarding $\widehat{\beta}_{B_k,j}$, i.e., the estimated marginal association statistic between $G_j$ and $B_k$: for $G_j = (G_{j,1}, \ldots, G_{j,N})$, i.e., the vector of $N$ observations of SNP $j$ that is selected as one of the genetic IVs for $B_k$, we have the following properties:

$$
\begin{aligned}
E(\widehat{\beta}_{B_k,j}|\beta_{x,j}, \gamma_{k,j}s) &= E\big[(\boldsymbol{G_j}^T\boldsymbol{G}_j)^{-1}\boldsymbol{G_j}^T\boldsymbol{B}_k|\beta_{x,j}, \gamma_{k,j}s\big] \\
&= E\Big[(\boldsymbol{G_j}^T\boldsymbol{G}_j)^{-1}\boldsymbol{G_j}^T(\theta_k\sum_{j=1}^{M}\beta_{x,j}\boldsymbol{G}_j + \sum_{j=1}^{M}\gamma_{k,j}\boldsymbol{G}_j + \theta_k\boldsymbol{\epsilon_x} + \boldsymbol{\epsilon_{Bk}})|\beta_{x,j}, \gamma_{k,j}s\Big] \\
&= \theta_k\beta_{x,j} + \gamma_{k,j}, \\
Var(\widehat{\beta}_{B_k,j}|\beta_{x,j}, \gamma_{k,j}s) &= Var\big[E\big((\boldsymbol{G_j}^T\boldsymbol{G}_j)^{-1}\boldsymbol{G_j}^T\boldsymbol{B}_k|\beta_{x,j}, \gamma_{k,j}s\big)\big] \\
&+ E\big[Var\big((\boldsymbol{G_j}^T\boldsymbol{G}_j)^{-1}\boldsymbol{G_j}^T\boldsymbol{B}_k|\beta_{x,j}, \gamma_{k,j}s\big)\big] \\
&= Var\big[\theta_k\beta_{x,j} + \gamma_{k,j}|\beta_{x,j}, \gamma_{k,j}s\big] \\
&+ E\big[(\boldsymbol{G_j}^T\boldsymbol{G}_j)^{-1}\boldsymbol{G_j}^T(\theta_k^2\sigma_{\epsilon_x}^2 + \sigma_{\epsilon_{B_k}}^2)\boldsymbol{I}\boldsymbol{G}_j(\boldsymbol{G_j}^T\boldsymbol{G}_j)^{-1}|\beta_{x,j}, \gamma_{k,j}s\big] \\
&= E\big[(\boldsymbol{G_j}^T\boldsymbol{G}_j)^{-1}(\theta_k^2\sigma_{\epsilon_x}^2 + \sigma_{\epsilon_{B_k}}^2)\big] \\
&= \frac{1}{n_{k,j}}(\theta_k^2\sigma_{\epsilon_x}^2 + \sigma_{\epsilon_{B_k}}^2), \\
Cov(\widehat{\beta}_{B_k,j}, \widehat{\beta}_{B_k,l}) &= Cov\big((\boldsymbol{G_j}^T\boldsymbol{G}_j)^{-1}\boldsymbol{G_j}^T(\theta_k\beta_{x,j} + \gamma_{k,j})\boldsymbol{G}_j, (\boldsymbol{G_l}^T\boldsymbol{G}_l)^{-1}\boldsymbol{G_l}^T(\theta_k\beta_{x,l} + \gamma_{k,l})\boldsymbol{G}_l\big) \\
&+ E\big[(\boldsymbol{G_j}^T\boldsymbol{G}_j)^{-1}\boldsymbol{G_j}^T(\theta_k^2\sigma_{\epsilon_x}^2 + \sigma_{\epsilon_{B_k}}^2)\boldsymbol{I}\boldsymbol{G}_l(\boldsymbol{G_l}^T\boldsymbol{G}_l)^{-1}\big] \\
&= \frac{n_{k,jl}d_{j,l}}{n_{k,j}n_{k,l}}(\theta_k^2\sigma_{\epsilon_x}^2 + \sigma_{\epsilon_{B_k}}^2) \\
&= 0,
\end{aligned}
$$

where $n_{k,j}$ and $d_{j,l}$ denote the sample overlap in the GWAS for $B_k$ and correlation, respectively, for SNP $j$ and SNP $l$, $l, j = 1, \ldots, M$ and $k = 1, \ldots, K$. Since we only consider independent IVs for now, $d_{j,l} = 0$. $\sqrt{\boldsymbol{n_k}}^T\left(\widehat{\boldsymbol{\beta}}_{B_k} - (\theta_k\boldsymbol{\beta}_x + \boldsymbol{\gamma}_k)\right) \xrightarrow{d} \mathcal{N}\left(\boldsymbol{0}, (\theta_k^2\sigma_{\epsilon_x}^2 + \sigma_{\epsilon_{B_k}}^2)\boldsymbol{I}\right)$, where $\boldsymbol{n_k} = (n_{k,1}, \ldots, n_{k,M})^T$. Similarly, for the estimated marginal association statistic between $G_j$ and $Y$ ($\widehat{\beta}_{Y,j}$), $j = 1, \ldots, M$:

$$\begin{aligned} E(\widehat{\beta}_{Y,j}) &= \theta\beta_{x,j}, \\ Var(\widehat{\beta}_{Y,j}) &= \frac{1}{n_{y,j}}(\theta^2\sigma^2_{\epsilon_x} + \sigma^2_{\epsilon_y}), \\ Cov(\widehat{\beta}_{Y,j}, \widehat{\beta}_{Y,l}) &= 0. \end{aligned}$$

With $n_{y,j} \to \infty$ and fixed $M$, $\sqrt{\boldsymbol{n_y}}^T \left(\widehat{\boldsymbol{\beta}}_Y - \theta\boldsymbol{\beta}_x\right) \xrightarrow{d} \mathcal{N}\left(\mathbf{0}, (\theta^2\sigma^2_{\epsilon_x} + \sigma^2_{\epsilon_y})\boldsymbol{I}\right)$, where $\boldsymbol{n_y} = (n_{y,1}, \ldots, n_{y,M})^T$. We allow sample overlap between GWAS for $B_k$s with $Cov(\boldsymbol{\beta_{B_k}}, \boldsymbol{\beta_{B_{k'}}}) = \boldsymbol{c_{k,k'}}$, $1 \leq k < k' \leq K$ that can be estimated from the intercept in bi-variate LD score regressions, but assume that the summary association statistics for $B_k$s and $Y$ are from independent GWAS data source, so that $Cov(\boldsymbol{\beta_Y}, \boldsymbol{\beta_{B_k}}) = \mathbf{0}$, $k = 1, \ldots, K$. We can derive the RSS likelihood of the summary date

$$\begin{aligned} & L_{rss}\left(\boldsymbol{\vartheta}; \{\widehat{\boldsymbol{\beta}}_{B_k}\}_{k=1}^K, \widehat{\boldsymbol{\beta}}_Y, \{\widehat{\boldsymbol{\sigma}}^2_{B_k}\}_{k=1}^K, \widehat{\boldsymbol{\sigma}}^2_Y\right) \\ = & \prod_{k=1}^K p\left(\widehat{\boldsymbol{\beta}}_{B_k} | \boldsymbol{\vartheta}, \widehat{\boldsymbol{\sigma}}^2_{B_k}\right) \times p\left(\widehat{\boldsymbol{\beta}}_Y | \boldsymbol{\vartheta}, \widehat{\boldsymbol{\sigma}}^2_Y\right) \\ = & \prod_{k=1}^K \prod_{j=1}^M N\left(\widehat{\beta}_{B_k,j} | \theta_k\beta_{x,j} + \gamma_{k,j},\ \frac{1}{n_k}(\theta_k^2\sigma^2_{\epsilon_x} + \sigma^2_{\epsilon_{B_k}})\right) \times \prod_{j=1}^M N\left(\widehat{\beta}_{Y,j} | \theta\beta_{x,j}, \frac{1}{n_y}(\theta^2\sigma^2_{\epsilon_x} + \sigma^2_{\epsilon_y})\right), \end{aligned}$$

where $\boldsymbol{\vartheta} = \left(\theta, \{\theta_k\}_{k=1}^K, \{\tau_k^2\}_{k=1}^K, \boldsymbol{\beta}_x, \boldsymbol{\gamma}_1, \ldots, \boldsymbol{\gamma}_K\right)^T$ denotes the set of parameters of interest. We then replace the covariance matrices with their sample estimates, and the RSS likelihood becomes:

$$
\begin{aligned}
& L_{rss}\left(\boldsymbol{\vartheta}; \{\widehat{\boldsymbol{\beta}}_{B_k}\}_{k=1}^{K}, \widehat{\boldsymbol{\beta}}_Y, \{\widehat{\boldsymbol{\sigma}}^2_{B_k}\}_{k=1}^{K}, \widehat{\boldsymbol{\sigma}}^2_Y\right) \\
= & \prod_{k=1}^{K}\prod_{j=1}^{M} N\left(\widehat{\beta}_{B_k,j} | \theta_k \beta_{x,j} + \gamma_{k,j}, \widehat{\sigma}^2_{B_k,j}\right) \times \prod_{j=1}^{M} N\left(\widehat{\beta}_{Y,j} | \theta \beta_{x,j}, \widehat{\sigma}^2_{Y,j}\right) \\
= & \mathcal{N}\Bigg( \begin{bmatrix} \theta_1 \boldsymbol{\beta}_x + \boldsymbol{\gamma}_1 \\ \cdots \\ \theta_K \boldsymbol{\beta}_x + \boldsymbol{\gamma}_K \\ \theta \boldsymbol{\beta}_x \end{bmatrix}, \begin{bmatrix} (\theta_1^2 \sigma_x^2 + \sigma_1^2)\text{diag}(n_1^{-1}) & \cdots & Cov(\widehat{\boldsymbol{\beta}}_{B_1}, \widehat{\boldsymbol{\beta}}_{B_K}) & \mathbf{0} \\ \cdots & \cdots & \cdots & \cdots \\ Cov(\widehat{\boldsymbol{\beta}}_{B_1}, \widehat{\boldsymbol{\beta}}_{B_K}) & \cdots & (\theta_K^2 \sigma_x^2 + \sigma_K^2)\text{diag}(n_K^{-1}) & \mathbf{0} \\ \mathbf{0} & \cdots & \mathbf{0} & (\theta^2 \sigma_x^2 + \sigma_y^2)\text{diag}(n_y^{-1}) \end{bmatrix} \Bigg) \\
= & \mathcal{N}\Bigg( \begin{bmatrix} \theta_1 \boldsymbol{\beta}_x + \boldsymbol{\gamma}_1 \\ \cdots \\ \theta_K \boldsymbol{\beta}_x + \boldsymbol{\gamma}_K \\ \theta \boldsymbol{\beta}_x \end{bmatrix}, \begin{bmatrix} \text{diag}(\widehat{\boldsymbol{\sigma}}^2_{B_1}) & \cdots & \boldsymbol{c}_{1,K} = \widehat{\rho}_{1,K} \widehat{\boldsymbol{\sigma}}_{B_1} \widehat{\boldsymbol{\sigma}}^T_{B_K} & \mathbf{0} \\ \cdots & \cdots & \cdots & \cdots \\ \boldsymbol{c}_{1,K} = \widehat{\rho}_{1,K} \widehat{\boldsymbol{\sigma}}_{B_1} \widehat{\boldsymbol{\sigma}}^T_{B_K} & \cdots & \text{diag}(\widehat{\boldsymbol{\sigma}}^2_{B_K}) & \mathbf{0} \\ \mathbf{0} & \cdots & \mathbf{0} & \text{diag}(\widehat{\boldsymbol{\sigma}}^2_Y) \end{bmatrix} \Bigg) \\
= & \mathcal{N}\Big( \boldsymbol{A}_\theta \boldsymbol{\beta}_x + \boldsymbol{B}_\gamma, \boldsymbol{\Omega}_{\sigma^2} \Big),
\end{aligned}
$$

where $\widehat{\rho}_{j,l}$ denotes the estimated genetic correlation between biomarker $B_j$ and $B_l$ from the intercept in bi-variate LD score regressions, $\boldsymbol{A}_\theta = [\text{diag}(\boldsymbol{\theta}_1), \cdots, \text{diag}(\boldsymbol{\theta}_K), \text{diag}(\boldsymbol{\theta})]^T$, and $\boldsymbol{B}_\gamma = [\boldsymbol{\gamma}_1, \cdots, \boldsymbol{\gamma}_K, \mathbf{0}]^T$. We can then derive the full conditional distributions of the model parameters.

Denote $\boldsymbol{S} = [\widehat{\boldsymbol{\beta}}_{B_1}, \cdots, \widehat{\boldsymbol{\beta}}_{B_K}, \widehat{\boldsymbol{\beta}}_Y]^T$ and assume the conjugate prior $\boldsymbol{\beta}_x \sim \mathcal{N}(\mathbf{0}, \text{diag}(\boldsymbol{\tau}_x^2))$, i.e., $\beta_{x,j} \sim \mathcal{N}(0, \tau_x^2)$, the full conditional distribution for $\boldsymbol{\beta}_x$ is

$$
\begin{aligned}
& \boldsymbol{\beta}_x | \boldsymbol{S} \propto \mathcal{N}\Big( \boldsymbol{S} | \boldsymbol{A}_\theta \boldsymbol{\beta}_x + \boldsymbol{B}_\gamma, \boldsymbol{\Omega}_{\sigma^2} \Big) \times \mathcal{N}\Big( \boldsymbol{\beta}_x | \mathbf{0}, \text{diag}(\boldsymbol{\tau}_x^2) \Big) \\
\propto & \ \mathcal{N}\Big( (\boldsymbol{A}_\theta^T \boldsymbol{A}_{\boldsymbol{\theta}})^{-1} \boldsymbol{A}_\theta^T \boldsymbol{S} - (\boldsymbol{A}_\theta^T \boldsymbol{A}_\theta)^{-1} \boldsymbol{A}_\theta^T \boldsymbol{B}_\gamma | \boldsymbol{\beta}_x, \boldsymbol{C} \Big) \times \mathcal{N}\Big( \boldsymbol{\beta}_x | \mathbf{0}, \text{diag}(\boldsymbol{\tau}_x^2) \Big) \\
\propto & \ \mathcal{N}\Big( \frac{\boldsymbol{C}^{-1}[(\boldsymbol{A}_\theta^T \boldsymbol{A}_\theta)^{-1} \boldsymbol{A}_\theta^T (\boldsymbol{S} - \boldsymbol{B}_\gamma)]}{\boldsymbol{C}^{-1} + \text{diag}(\boldsymbol{\tau}_x^{-2})}, \Big[ \boldsymbol{C}^{-1} + \text{diag}(\boldsymbol{\tau}_x^{-2}) \Big]^{-1} \Big),
\end{aligned}
$$

where $\boldsymbol{C} = (\boldsymbol{A}_\theta^T \boldsymbol{A}_\theta)^{-1} \boldsymbol{A}_\theta^T \boldsymbol{\Omega}_{\sigma^2} \boldsymbol{A}_\theta (\boldsymbol{A}_\theta^T \boldsymbol{A}_\theta)^{-1}$. Notice in the MRLE paper by Jin et al. (2024), there is a potential convergence issue at $h_x^2$ since it always appear alongside $\theta$, $\theta_k$, and $\theta\theta_k$. In CaLMR (Uni), we fix the prior variance of $\boldsymbol{\beta}_x$ to address the issue. The second step is updating the first $K \times M$ non-zero entries of $\boldsymbol{B}_\gamma$. For IV $G_j$, denote $\boldsymbol{\gamma}_j = (\gamma_{1,j}, \gamma_{2,j}, \cdots, \gamma_{K,j})^T$, $\boldsymbol{S}_j = (\hat{\beta}_{B_1,j}, \cdots, \hat{\beta}_{B_K,j})^T$, and the corresponding covariance matrix $\boldsymbol{\Omega}_j = \begin{bmatrix} \hat{\sigma}_{1,j}^2 & \cdots & \hat{\rho}_{1,K}\hat{\sigma}_{1,j}\hat{\sigma}_{K,j} \\ \cdots & \cdots & \cdots \\ \hat{\rho}_{1,K}\hat{\sigma}_{1,j}\hat{\sigma}_{K,j} & \cdots & \hat{\sigma}_{K,j}^2 \end{bmatrix}$. With conjugate prior $\boldsymbol{\gamma}_j \sim \mathcal{N}\Big(\mathbf{0}, \text{diag}(\tau_1^2, \dots, \tau_K^2)\Big)$, the full conditional for $\boldsymbol{\gamma}_j$ is then:

$$
\begin{aligned}
\boldsymbol{\gamma}_j | \boldsymbol{S}_j &\propto \mathcal{N}(\boldsymbol{S}_j | (\theta_1, \dots, \theta_K)^T \beta_{x,j} + \boldsymbol{\gamma}_j, \boldsymbol{\Omega}_j) \times \mathcal{N}(\boldsymbol{\gamma}_j | \mathbf{0}, \text{diag}(\tau_1^2, \dots, \tau_K^2)) \\
&\propto \mathcal{N}\Big(\boldsymbol{S}_j - (\theta_1, \dots, \theta_K)^T \beta_{x,j} | \boldsymbol{\gamma}_j, \boldsymbol{\Omega}_j\Big) \times \mathcal{N}(\boldsymbol{\gamma}_j | \mathbf{0}, \text{diag}(\tau_1^2, \dots, \tau_K^2)) \\
&\propto \mathcal{N}\Big(\frac{\boldsymbol{\Omega}_j^{-1}[\boldsymbol{S}_j - (\theta_1, \dots, \theta_K)^T \beta_{x,j}]}{\boldsymbol{\Omega}_j^{-1} + \text{diag}(\tau_1^{-2}, \dots, \tau_K^{-2})}, \Big[\boldsymbol{\Omega}_j^{-1} + \text{diag}(\tau_1^{-2}, \dots, \tau_K^{-2})\Big]^{-1}\Big).
\end{aligned}
$$

We update the prior variance, $\{\tau_k^2\}_{k=1}^K$, by assuming an Inverse-Gamma prior $\tau_k^2 \sim \mathcal{IG}(\alpha_k^0, \beta_k^0)$, so that the full conditional distribution for $\tau_k^2$

$$
\begin{aligned}
\tau_k^2 | \boldsymbol{S} &\propto \mathcal{IG}(\tau_k^2 | \alpha_k^0, \beta_k^0) \times \mathcal{N}\Big(\boldsymbol{\gamma}_k | \mathbf{0}, \text{diag}(\boldsymbol{\tau}_k^2)\Big) \\
&\propto \mathcal{IG}\Big(\alpha_k^0 + \frac{M}{2}, \beta_k^0 + \frac{1}{2}\sum_{j=1}^{M} \gamma_{k,j}^2\Big).
\end{aligned}
$$

The final step is updating $\boldsymbol{\eta}_\theta = (\theta_1, \cdots, \theta_K, \theta)^T$ with a non-informative prior, so that the sampling of $\boldsymbol{\eta}_\theta$ completely depends on the summary data. The full conditional distribution for $\boldsymbol{\eta}_\theta$ is then

$$\boldsymbol{\eta}_\theta \mid \boldsymbol{S} \;\propto\; \prod_{j=1}^{M} \mathcal{N}\Big( \begin{bmatrix} \hat{\beta}_{B_1,j} \\ \cdots \\ \hat{\beta}_{B_K,j} \\ \hat{\beta}_{Y,j} \end{bmatrix} \mid \begin{bmatrix} \theta_1\beta_{x,j} + \gamma_{1,j} \\ \cdots \\ \theta_K\beta_{x,j} + \gamma_{K,j} \\ \theta\beta_{x,j} + 0 \end{bmatrix}, \boldsymbol{\Delta}_j = \begin{bmatrix} \hat{\sigma}^2_{1,j} & \cdots & \hat{\rho}_{1,K}\hat{\sigma}_{1,j}\hat{\sigma}_{K,j} & 0 \\ \cdots & \cdots & \cdots & \cdots \\ \hat{\rho}_{1,K}\hat{\sigma}_{1,j}\hat{\sigma}_{K,j} & \cdots & \hat{\sigma}^2_{K,j} & \cdots \\ 0 & \cdots & \cdots & \hat{\sigma}^2_{Y,j} \end{bmatrix} \Big)$$

$$\propto \prod_{j=1}^{M} \mathcal{N}\Big( \boldsymbol{S}_j | \beta_{x,j}\boldsymbol{\eta}_\theta + \boldsymbol{\gamma}_j, \boldsymbol{\Delta}_j \Big)$$

$$\propto \prod_{j=1}^{M} \mathcal{N}\Big( \beta_{x,j}^{-1}(\boldsymbol{S}_j - \boldsymbol{\gamma}_j) | \boldsymbol{\eta}_\theta, \beta_{x,j}^{-2}\boldsymbol{\Delta}_j \Big)$$

$$\propto \mathcal{N}\Big( \left[ \sum_{j=1}^{M} (\beta_{x,j}^{-2}\boldsymbol{\Delta}_j)^{-1} \right]^{-1} \left[ \sum_{j=1}^{M} (\beta_{x,j}^{-2}\boldsymbol{\Delta}_j)^{-1}\beta_{x,j}^{-1}(\boldsymbol{S}_j - \boldsymbol{\gamma}_j) \right], \left[ \sum_{j=1}^{M} (\beta_{x,j}^{-2}\boldsymbol{\Delta}_j)^{-1} \right]^{-1} \Big).$$

#### 1.2 Multivariate Case: Multiple Latent Exposures

When there are multiple correlated latent exposures presenting, recall the random effects models are:

$$\begin{aligned} Y &= \sum_{l=1}^{L} \theta_l X_l + \epsilon_y, \quad \epsilon_y \sim N(0, \sigma^2_{\epsilon_y}) \\ X_l &= \sum_{j=1}^{M} \beta_{x_l,j} G_j + \epsilon_{x_l}, \quad \epsilon_{x_l} \sim N(0, \sigma^2_{\epsilon_{x_l}}) \\ B_k &= \sum_{l=1}^{L} \theta_{kl} X_l + \sum_{j=1}^{M} \gamma_{k,j} G_j + \epsilon_{B_k}, \quad \epsilon_{B_k} \sim N(0, \sigma^2_{\epsilon_{B_k}}), \end{aligned}$$

where $\beta_{x_l,j} \sim N(0, h^2_{x_l,j})$ and $\gamma_{k,j} \sim N(0, h^2_{\gamma_k})$. Like we did in the univariate scenario, the estimated marginal association statistic between $G_j$ and $B_k$, $\widehat{\beta})B_k, j$ has the following results:

$$
\begin{aligned}
E(\widehat{\beta}_{B_k,j}|\beta_{x_l,j},\gamma_{k,j}s) &= E\big[(\boldsymbol{G}_j{}^T\boldsymbol{G}_j)^{-1}\boldsymbol{G}_j{}^T\boldsymbol{B}_k|\beta_{x_l,j},\gamma_{k,j}s\big] \\
&= E\Big[(\boldsymbol{G}_j{}^T\boldsymbol{G}_j)^{-1}\boldsymbol{G}_j{}^T(\sum_{l=1}^{L}\sum_{j=1}^{M}(\theta_{kl}\beta_{x_l,j}+\gamma_{k,j})\boldsymbol{G}_j+\sum_{l=1}^{L}\theta_{kl}\boldsymbol{\epsilon}_{\boldsymbol{x_l}}+\boldsymbol{\epsilon}_{\boldsymbol{B_k}})|\beta_{x_l,j},\gamma_{k,j}s\Big] \\
&= E\Big[\theta_{k1}\beta_{x_1,j}+\ldots+\theta_{kL}\beta_{x_L,j}+\gamma_{k,j}\Big] \\
&= \sum_{l=1}^{L}\theta_{kl}\beta_{x_l,j}+\gamma_{k,j}, \\
Var(\widehat{\beta}_{B_k,j}|\beta_{x_l,j},\gamma_{k,j}s) &= Var\big[E\big((\boldsymbol{G}_j{}^T\boldsymbol{G}_j)^{-1}\boldsymbol{G}_j{}^T\boldsymbol{B}_k|\beta_{x_l,j},\gamma_{k,j}s\big)\big]+E\big[Var\big((\boldsymbol{G}_j{}^T\boldsymbol{G}_j)^{-1}\boldsymbol{G}_j{}^T\boldsymbol{B}_k|\beta_{x_l,j},\gamma_{k,j} \\
&= Var\big[\sum_{l=1}^{L}\theta_{kl}\beta_{x_l,j}+\gamma_{k,j}|\beta_{x_l,j},\gamma_{k,j}s\big] \\
&+ E\big[(\boldsymbol{G}_j{}^T\boldsymbol{G}_j)^{-1}\boldsymbol{G}_j{}^T(\sum_{l=1}^{L}\theta_{kl}^2\sigma_{\epsilon_{x_l}}^2+\sigma_{\epsilon_{B_k}}^2)\boldsymbol{I}\boldsymbol{G}_j(\boldsymbol{G}_j{}^T\boldsymbol{G}_j)^{-1}|\beta_{x_l,j},\gamma_{k,j}s\big] \\
&= E\big[(\boldsymbol{G}_j{}^T\boldsymbol{G}_j)^{-1}(\sum_{l=1}^{L}\theta_{kl}^2\sigma_{\epsilon_{x_l}}^2+\sigma_{\epsilon_{B_k}}^2)\big] \\
&= \frac{1}{n_{k,j}}(\sum_{l=1}^{L}\theta_{kl}^2\sigma_{\epsilon_{x_l}}^2+\sigma_{\epsilon_{B_k}}^2),
\end{aligned}
$$

$$
\begin{aligned}
Cov(\widehat{\beta}_{B_k,i},\widehat{\beta}_{B_k,j}) &= Cov\big((\boldsymbol{G}_i{}^T\boldsymbol{G}_i)^{-1}\boldsymbol{G}_i{}^T(\sum_{l=1}^{L}\theta_{kl}\beta_{x_l,i}+\gamma_{k,i})\boldsymbol{G}_i,(\boldsymbol{G}_j{}^T\boldsymbol{G}_j)^{-1}\boldsymbol{G}_j{}^T(\sum_{l=1}^{L}\theta_{kl}\beta_{x_l,j}+\gamma_{k,j})\boldsymbol{G}_j\big) \\
&+ E\big[(\boldsymbol{G}_i{}^T\boldsymbol{G}_i)^{-1}\boldsymbol{G}_i{}^T(\theta_{kl}^2\sigma_{\epsilon_{x_l}}^2+\sigma_{\epsilon_{B_k}}^2)\boldsymbol{I}\boldsymbol{G}_j(\boldsymbol{G}_j{}^T\boldsymbol{G}_j)^{-1}\big] \\
&= \frac{n_{k,ij}d_{i,j}}{n_{k,i}n_{k,j}}(\sum_{l=1}^{L}\theta_{kl}^2\sigma_{\epsilon_{x_l}}^2+\sigma_{\epsilon_{B_k}}^2)=0,
\end{aligned}
$$

where $n_{k,ij}$ and $d_{i,j}$ denote the sample overlap in the GWAS for $B_k$ and correlation, respectively, for SNP $i$ and SNP $j$, with $i,j=1,\ldots,M$ and $k=1,\ldots,K$. Since we still only consider independent IVs, $d_{i,j}=0$. For $\widehat{\beta}_{Y,j}$, the estimated marginal association statistic between $G_j$ and $Y$, $j = 1, \dots, M$:

$$\begin{aligned} E(\widehat{\beta}_{Y,j}) &= \sum_{l=1}^{L} \theta_l \beta_{x_l,j}, \\ Var(\widehat{\beta}_{Y,j}) &= \frac{1}{n_{y,j}} (\sum_{l=1}^{L} \theta_l^2 \sigma^2_{\epsilon_{x_l}} + \sigma^2_{\epsilon_y}), \\ Cov(\widehat{\beta}_{Y,j}, \widehat{\beta}_{Y,l}) &= 0. \end{aligned}$$

With $n_{y,j} \to \infty$ and finite $M$, $\sqrt{\boldsymbol{n_y}}^T \left( \widehat{\boldsymbol{\beta}}_Y - \sum_{l=1}^{L} \theta_l \boldsymbol{\beta}_{x_l} \right) \xrightarrow{d} \mathcal{N}\left( \mathbf{0}, (\sum_{l=1}^{L} \theta_l^2 \sigma^2_{\epsilon_{x_l}} + \sigma^2_{\epsilon_y}) \boldsymbol{I} \right)$, where $\boldsymbol{n_y} = (n_{y,1}, \dots, n_{y,M})^T$. In the multiple-exposure setting, we also allow sample overlap between GWAS for various observable traits $B_k$s but the summary association statistics for $B_k$s and $Y$ are from independent data sources, so that $Cov(\boldsymbol{\beta_{B_k}}, \boldsymbol{\beta_{B_{k'}}}) = c_{k,k'}$, $1 \le k < k' \le K$ and $Cov(\boldsymbol{\beta_Y}, \boldsymbol{\beta_{B_k}}) = \mathbf{0}$, $k = 1, \dots, K$. These conditional expectations and conditional variances results help us to extend the RSS likelihood to the multivariate MR setting:

$$\begin{aligned} & L_{rss}(\boldsymbol{\vartheta}; \{\widehat{\boldsymbol{\beta}}_{B_k}\}, \widehat{\boldsymbol{\beta}}_Y, \{\widehat{\boldsymbol{\sigma^2}_{B_k}}\}, \widehat{\boldsymbol{\sigma^2}_Y}) \\ = & \mathcal{N}\Bigg( \begin{bmatrix} \theta_{11}\boldsymbol{\beta}_{x_1} + \ldots + \theta_{1L}\boldsymbol{\beta}_{x_L} + \boldsymbol{\gamma}_1 \\ \theta_{21}\boldsymbol{\beta}_{x_1} + \ldots + \theta_{2L}\boldsymbol{\beta}_{x_L} + \boldsymbol{\gamma}_2 \\ \cdots \\ \theta_{K1}\boldsymbol{\beta}_{x_1} + \ldots + \theta_{KL}\boldsymbol{\beta}_{x_L} + \boldsymbol{\gamma}_K \\ \theta_1\boldsymbol{\beta}_{x_1} + \ldots + \theta_L\boldsymbol{\beta}_{x_L} \end{bmatrix}, \boldsymbol{\Omega}_{\sigma^2} = \begin{bmatrix} \mathrm{diag}(\widehat{\boldsymbol{\sigma}}_1^2) & \cdots & \boldsymbol{c}_{1,K} & \mathbf{0} \\ \cdots & \cdots & \cdots & \cdots \\ \boldsymbol{c}_{1,K} & \cdots & \mathrm{diag}(\widehat{\boldsymbol{\sigma}}_K^2) & \mathbf{0} \\ \mathbf{0} & \cdots & \mathbf{0} & \mathrm{diag}(\widehat{\boldsymbol{\sigma}}_Y^2). \end{bmatrix} \Bigg) \\ = & \mathcal{N}\Big( \sum_{l=1}^{L} \boldsymbol{A}_l \boldsymbol{\beta}_{x_l} + \boldsymbol{B}_\gamma, \boldsymbol{\Omega}_{\sigma^2} \Big), \end{aligned}$$

where $\boldsymbol{A}_l = [\mathrm{diag}(\boldsymbol{\theta}_{1l}), \cdots, \mathrm{diag}(\boldsymbol{\theta}_{Kl}), \mathrm{diag}(\boldsymbol{\theta}_l)]^T$ and $\boldsymbol{B}_\gamma = [\boldsymbol{\gamma}_1, \cdots, \boldsymbol{\gamma}_K, \mathbf{0}]^T$.

In the MCMC algorithm, we first assume a conjugate prior $\boldsymbol{\beta}_j = (\beta_{x_1,j}, \dots, \beta_{x_L,j})^T \sim \mathcal{N}(\mathbf{0}, \boldsymbol{H})$, where $\boldsymbol{H} = \boldsymbol{DRD}$. $\boldsymbol{R}$ denotes the correlation matrix for the latent exposures and $\boldsymbol{D} = \text{diag}(h_{x_1}, \ldots, h_{x_L})$. Let $\boldsymbol{S}_j = (\hat{\beta}_{B_1,j}, \ldots, \hat{\beta}_{B_K,j}, \hat{\beta}_{Y,j})^T$ be a vector of the summary statistics for IV $G_j$ and $\Delta_j = \begin{bmatrix} \hat{\sigma}_{1,j} & \ldots & \hat{\rho}_{1,K}\hat{\sigma}_{1,j}\hat{\sigma}_{K,j} & 0 \\ \ldots & \ldots & \ldots & \ldots \\ \hat{\rho}_{1,K}\hat{\sigma}_{1,j}\hat{\sigma}_{K,j} & \ldots & \hat{\sigma}_{K,j} & 0 \\ 0 & \ldots & 0 & \hat{\sigma}_{Y,j} \end{bmatrix}$ be the corresponding covariance matrix. The full conditional distribution for $\boldsymbol{\beta}_j$:

$$
\begin{aligned}
\boldsymbol{\beta}_j \mid \boldsymbol{S} \;&\propto\; \mathcal{N}\Big(\boldsymbol{S}_j | \begin{bmatrix} \theta_{11}\boldsymbol{\beta}_{x_1,j} + \ldots + \theta_{1L}\boldsymbol{\beta}_{x_L,j} + \boldsymbol{\gamma}_{1,j} \\ \ldots \\ \theta_{K1}\boldsymbol{\beta}_{x_1,j} + \ldots + \theta_{KL}\boldsymbol{\beta}_{x_L,j} + \boldsymbol{\gamma}_{K,j} \\ \theta_1\boldsymbol{\beta}_{x_1,j} + \ldots + \theta_L\boldsymbol{\beta}_{x_L,j} \end{bmatrix}, \boldsymbol{\Delta}_j\Big) \times \mathcal{N}(\boldsymbol{\beta}_j | \boldsymbol{0}, \boldsymbol{H}) \\
&\propto\; \mathcal{N}\Big(\boldsymbol{S}_j | \begin{bmatrix} \theta_{11} & \ldots & \theta_{1L} \\ \ldots & \ldots & \ldots \\ \theta_{K1} & \ldots & \theta_{KL} \\ \theta_1 & \ldots & \theta_L \end{bmatrix} (\beta_{x_1,j}, \cdots, \beta_{x_L,j})^T + \begin{bmatrix} \gamma_{1,j} \\ \ldots \\ \gamma_{K,j} \\ 0 \end{bmatrix}, \boldsymbol{\Delta}_j\Big) \times \mathcal{N}(\boldsymbol{\beta}_j | \boldsymbol{0}, \boldsymbol{H}) \\
&\propto\; \mathcal{N}\Big(\boldsymbol{S}_j | \boldsymbol{A}_\theta \boldsymbol{\beta}_j + \boldsymbol{\gamma}_j, \boldsymbol{\Delta}_j\Big) \times \mathcal{N}(\boldsymbol{\beta}_j | \boldsymbol{0}, \boldsymbol{H}) \\
&\propto\; \mathcal{N}\Big((\boldsymbol{A}_\theta^T \boldsymbol{A}_\theta)^{-1} \boldsymbol{A}_\theta^T (\boldsymbol{S}_j - \boldsymbol{\gamma}_j) | \boldsymbol{\beta}_j, (\boldsymbol{A}_\theta^T \boldsymbol{A}_\theta)^{-1} \boldsymbol{A}_\theta^T \boldsymbol{\Delta}_j \boldsymbol{A}_\theta (\boldsymbol{A}_\theta^T \boldsymbol{A}_\theta)^{-1}\Big) \times \mathcal{N}(\boldsymbol{\beta}_j | \boldsymbol{0}, \boldsymbol{H}) \\
&\propto\; \mathcal{N}\Big(\Big[(\boldsymbol{C}_j^{-1} + \boldsymbol{H}^{-1})\Big]^{-1} \Big[\boldsymbol{C}_j^{-1}[\boldsymbol{A}_\theta^T \boldsymbol{A}_\theta)^{-1} \boldsymbol{A}_\theta^T (\boldsymbol{S}_j - \boldsymbol{\gamma}_j)]\Big], \Big[(\boldsymbol{C}_j^{-1} + \boldsymbol{H}^{-1})\Big]^{-1}\Big),
\end{aligned}
$$

where $\boldsymbol{C}_j = (\boldsymbol{A}_\theta^T \boldsymbol{A}_\theta)^{-1} \boldsymbol{A}_\theta^T \boldsymbol{\Delta}_j \boldsymbol{A}_\theta (\boldsymbol{A}_\theta^T \boldsymbol{A}_\theta)^{-1}$. The between-exposure correlation matrix, $\boldsymbol{R}$, is updated by first generating the posterior samples of $\boldsymbol{H}$ using an Inverse-Wishart prior $\mathcal{W}^{-1}(\boldsymbol{\Psi} = \boldsymbol{D}\boldsymbol{D}, \nu = L + 1)$. The full conditional distribution for $\boldsymbol{H}$ is then

$$
\begin{aligned}
\boldsymbol{H}' | \boldsymbol{S} \;&\propto\; \mathcal{W}^{-1}(\boldsymbol{H} | \boldsymbol{\Psi} = \boldsymbol{D}\boldsymbol{D}, \nu = L + 1) \times \mathcal{N}(\boldsymbol{\beta}_j | \boldsymbol{0}, \boldsymbol{H}) \\
&\propto\; \mathcal{W}^{-1}(\boldsymbol{\beta}_x \boldsymbol{\beta}_x^T + \boldsymbol{\Psi}, M + \nu).
\end{aligned}
$$

We calculate $\tilde{\boldsymbol{R}} = \boldsymbol{D}^{-1}\boldsymbol{H}^{'}\boldsymbol{D}^{-1}$ to get the posterior correlations. Notice we do not use $\boldsymbol{H}^{'}$ directly to update the prior covariance for $\boldsymbol{\beta}_j$. Instead, we plug in the posterior correlation $\tilde{\boldsymbol{R}}$ back to $\boldsymbol{H} = \boldsymbol{D}\tilde{\boldsymbol{R}}\boldsymbol{D}$, so that the prior variances of $\boldsymbol{\beta}_j$ is fixed to address the potential convergence issues, i.e., only the off-diagonal entries of $\boldsymbol{H}$ will be updated. Then, for IV $G_j$, denote $\boldsymbol{\gamma}_j = (\gamma_{1,j}, \gamma_{2,j}, \cdots, \gamma_{K,j})^T$, $\boldsymbol{S}_j = (\hat{\beta}_{B_1,j}, \cdots, \hat{\beta}_{B_K,j})^T$, and $\boldsymbol{\Omega}_j = \begin{bmatrix} \hat{\sigma}^2_{1,j} & \cdots & \hat{\rho}_{1,K}\hat{\sigma}_{1,j}\hat{\sigma}_{K,j} \\ \cdots & \cdots & \cdots \\ \hat{\rho}_{1,K}\hat{\sigma}_{1,j}\hat{\sigma}_{K,j} & \cdots & \hat{\sigma}^2_{K,j} \end{bmatrix}$. Assume the prior for $\boldsymbol{\gamma}_j$ is $\mathcal{N}\Big(\mathbf{0}, \boldsymbol{\Sigma} = \text{diag}(\tau_1^2, \ldots, \tau_K^2)\Big)$, the full conditional distribution for $\boldsymbol{\gamma}_j$

$$\begin{aligned}
\boldsymbol{\gamma}_j|\boldsymbol{S}_j &\propto \mathcal{N}(\boldsymbol{S}_j|\sum_{l=1}^{L}(\theta_{1l}, \ldots, \theta_{Kl})^T\beta_{x_l,j} + \boldsymbol{\gamma}_j, \boldsymbol{\Omega}_j) \times \mathcal{N}(\boldsymbol{\gamma}_j|\mathbf{0}, \boldsymbol{\Sigma}) \\
&\propto \mathcal{N}\Big(\boldsymbol{S}_j - \sum_{l=1}^{L}(\theta_{1l}, \ldots, \theta_{Kl})^T\beta_{x_l,j}|\boldsymbol{\gamma}_j, \boldsymbol{\Omega}_j\Big) \times \mathcal{N}(\boldsymbol{\gamma}_j|\mathbf{0}, \boldsymbol{\Sigma}) \\
&\propto \mathcal{N}\Big(\frac{\boldsymbol{\Omega}_j^{-1}[\boldsymbol{S}_j - \sum_{l=1}^{L}(\theta_{1l}, \ldots, \theta_{Kl})^T\beta_{x_l,j}]}{\boldsymbol{\Omega}_j^{-1} + \text{diag}(\tau_1^{-2}, \ldots, \tau_K^{-2})}, \Big[\boldsymbol{\Omega}_j^{-1} + \text{diag}(\tau_1^{-2}, \ldots, \tau_K^{-2})\Big]^{-1}\Big).
\end{aligned}$$

The prior variances of $\boldsymbol{B}_\gamma$ is updated by using an Inverse-gamma prior $\tau_k^2 \sim \mathcal{IG}(\alpha_k^0, \beta_k^0)$ to get the full conditional distribution

$$\begin{aligned}
\tau_k^2|\boldsymbol{S} &\propto \mathcal{IG}(\tau_k^2|\alpha_k^0, \beta_k^0) \times \mathcal{N}\Big(\gamma_k|\mathbf{0}, \text{diag}(\boldsymbol{\tau}_k^2)\Big) \\
&\propto \mathcal{IG}\Big(\alpha_k^0 + \frac{M}{2}, \beta_k^0 + \frac{1}{2}\sum_{j=1}^{M}\gamma_{k,j}^2\Big).
\end{aligned}$$

Finally, assume $B_l = \{B_{l1}, \ldots, B_{lm}\}$ is a vector containing the $m$ (i.e., $K_{x_l} = m$) biomarkers co-regulated by the latent exposure $X_l, l = 1, \ldots, L$ and we will update $\boldsymbol{\eta}_{\theta_l} = (\theta_{B_{l1}l}, \cdots, \theta_{B_{lm}l}, \theta_l)^T$ using a non-informative prior. Define the IV $G_j$ related summary statistics $\boldsymbol{S}_{lj} = (\hat{\beta}_{B_{l1},j}, \cdots, \hat{\beta}_{B_{lm},j}, \hat{\beta}_{Y,j})$

and their corresponding covariance matrix $\boldsymbol{\Delta}_j = \begin{bmatrix} \hat{\sigma}^2_{B_{l1},j} & \cdots & \hat{\rho}_{B_{l1},B_{lm}}\hat{\sigma}_{B_{l1},j}\hat{\sigma}_{B_{lm},j} & 0 \\ \cdots & \cdots & \cdots & \cdots \\ \hat{\rho}_{B_{l1},B_{lm}}\hat{\sigma}_{B_{l1},j}\hat{\sigma}_{B_{lm},j} & \cdots & \hat{\sigma}^2_{B_{lm},j} & 0 \\ 0 & \cdots & 0 & \hat{\sigma}^2_{Y,j} \end{bmatrix}$.

The full conditional distribution for $\boldsymbol{\eta}_{\theta_l}$ is

$$\begin{aligned}\boldsymbol{\eta}_{\theta_l} \mid \boldsymbol{S}_{lj} \;&\propto\; \prod_{j=1}^{M} \mathcal{N}\Big(\boldsymbol{S}_{lj} | \sum_{l=1}^{L} (\theta_{B_{l1}l}, \ldots, \theta_{B_{lm}l})^T \beta_{x_l,j} + \boldsymbol{\gamma}_{lj}), \boldsymbol{\Delta}_j\Big) \\ \propto\; \prod_{j=1}^{M} &\mathcal{N}\Big(\beta^{-1}_{x_l,j}[\boldsymbol{S}_{lj} - \sum_{i \neq l} (\theta_{B_{l1}i}, \ldots, \theta_{B_{lm}i})^T \beta_{x_i,j} - \boldsymbol{\gamma}_{lj}] | \boldsymbol{\eta}_{\theta_l}, \beta^{-2}_{x_l,j}\boldsymbol{\Delta}_j\Big) \\ \propto\; \mathcal{N}\Big(&\Big[\sum_{j=1}^{M} \beta^{-2}_{x_l,j}\boldsymbol{\Delta}_j\Big]^{-1} \Big[\sum_{j=1}^{M} (\frac{\boldsymbol{\Delta}_j}{\beta^2_{x_l,j}})^{-1} \frac{[\boldsymbol{S}_{lj} - \sum_{i \neq l}(\theta_{B_{l1}i}, \ldots, \theta_{B_{lm}i})^T \beta_{x_i,j} - \boldsymbol{\gamma}_{lj}]}{\beta_{x_l,j}}\Big], \Big[\sum_{j=1}^{M} \beta^{-2}_{x_l,j}\boldsymbol{\Delta}_j\Big]^{-1}\Big).\end{aligned}$$

#### 1.3 Identifiability Analysis

Since the prior variances of $\boldsymbol{\beta}_{x_l}$s are fixed, CaLMR can give inference to the significances and the directions of $\theta_l$s instead of the effect sizes, and the signs of $\theta_{kl}$s should be known to help identify the direction of causal effects. Specifically, if the sign of the posterior mean, $\bar{\theta}_{kl}$, is the opposite of its expected direction, we multiply the samples of $\theta_l$ and $\theta_{kl}, l = 1, \ldots, L, k = 1, \ldots, K$ by $-1$ to correct their directions. In the simulation studies, we observed the directions of the causal effects can be determined with 100 % accuracy as long as the directions of $\theta_{kl}s$ are pre-specified. However, our CaLMR test is still applicable even without prior knowledge of the signs of $\theta_{kl}$s. We should then be careful when interpreting the output results, since the directions of $\theta_l$s may be the opposite of what it should be. Also, sometimes the sign of the estimated between-exposure correlation $\boldsymbol{R}$ can be the opposite what it is, but the magnitude of the correlation is still valid.

### 2 Additional Simulation Results

#### 2.1 Simulation Results Under the Single Exposure Setting

In the simulation study with single latent exposure, we assumed there are $K = 6$ total observable traits. In all scenarios, CaLMR (Uni) reached 100 % accuracy in determining the direction of causal effects. The details of the single-exposure simulation results are as below.

Supplementary Table 1: Results summarized from 1000 simulations under the single exposure setting, with $K = 6$ total biomarkers. $\boldsymbol{\theta} = \mathbf{0/0.1}$, $H^2_{B_k} = \mathbf{0.2}$, and $H^2_x = 0.2$. The significance threshold for the IV selection is set at $\mathbf{5 \times 10^{-4}}$.

| $\theta$ | $N$ | $M$ | Method | Reject(%) | Time(m) |
|---|---|---|---|---|---|
| 0 | $6 \times 10^4$ | 67 | CaLMR (Uni) | 6.1 | 1.81 |
| | | | MRLE | 5.0 | 0.46 |
| | | | MR-PRESSO | 5.1 | 4.29 |
| | | | MR-IVW | 4.7 | - |
| | $8 \times 10^4$ | 113 | CaLMR (Uni) | 4.9 | 6.61 |
| | | | MRLE | 4.1 | 0.39 |
| | | | MR-PRESSO | 4.9 | 7.25 |
| | | | MR-IVW | 4.7 | - |
| | $1 \times 10^5$ | 165 | CaLMR (Uni) | 5.7 | 16.85 |
| | | | MRLE | 4.8 | 0.36 |
| | | | MR-PRESSO | 4.1 | 10.31 |
| | | | MR-IVW | 4.7 | - |
| 0.1 | $6 \times 10^4$ | 67 | CaLMR (Uni) | 78.4 | 1.81 |
| | | | MRLE | 83.4 | 0.43 |
| | | | MR-PRESSO | 7.7 | 4.04 |
| | | | MR-IVW | 7.8 | - |
| | $8 \times 10^4$ | 113 | CaLMR (Uni) | 99.3 | 6.45 |
| | | | MRLE | 99.6 | 0.39 |
| | | | MR-PRESSO | 9.3 | 7.02 |
| | | | MR-IVW | 10.0 | - |
| | $1 \times 10^5$ | 165 | CaLMR (Uni) | 100 | 17.25 |
| | | | MRLE | 99.9 | 0.40 |
| | | | MR-PRESSO | 12.3 | 10.07 |
| | | | MR-IVW | 12.7 | - |

"M" is the average number of IVs selected across 1000 simulations. "Reject(%)" denotes the percentage of the simulations where $H_0 : \theta = 0$ is rejected (i.e., type I error rate or power in this scenario), and "Time(m)" is measured in minutes.

Supplementary Table 2: Results summarized from 1000 simulations under the single exposure setting, with $K = 6$ total biomarkers. $\boldsymbol{\theta = 0/0.1}$, $H^2_{B_k} = \mathbf{0.3}$, and $H^2_x = 0.2$. The significance threshold for the IV selection is set at $\mathbf{5 \times 10^{-4}}$.

| $\theta$ | $N$ | $M$ | Method | Reject(%) | Time(m) |
|---|---|---|---|---|---|
| 0 | $6 \times 10^4$ | 139 | CaLMR (Uni) | 4.9 | 11.08 |
| | | | MRLE | 4.6 | 0.96 |
| | | | MR-PRESSO | 4.8 | 8.49 |
| | | | MR-IVW | 4.4 | - |
| | $8 \times 10^4$ | 177 | CaLMR (Uni) | 5.0 | 39.41 |
| | | | MRLE | 4.6 | 0.35 |
| | | | MR-PRESSO | 4.8 | 13.56 |
| | | | MR-IVW | 4.8 | - |
| | $1 \times 10^5$ | 246 | CaLMR (Uni) | 5.6 | 109.28 |
| | | | MRLE | 5.6 | 0.33 |
| | | | MR-PRESSO | 5.0 | 21.89 |
| | | | MR-IVW | 5.1 | - |
| 0.1 | $6 \times 10^4$ | 139 | CaLMR (Uni) | 100 | 11.53 |
| | | | MRLE | 99.9 | 0.39 |
| | | | MR-PRESSO | 12.3 | 9.69 |
| | | | MR-IVW | 13.3 | - |
| | $8 \times 10^4$ | 177 | CaLMR (Uni) | 100 | 40.43 |
| | | | MRLE | 99.8 | 0.40 |
| | | | MR-PRESSO | 17.4 | 16.20 |
| | | | MR-IVW | 18.1 | - |
| | $1 \times 10^5$ | 246 | CaLMR (Uni) | 100 | 98.95 |
| | | | MRLE | 100 | 0.42 |
| | | | MR-PRESSO | 24.3 | 21.94 |
| | | | MR-IVW | 24.8 | - |

"M" is the average number of IVs selected across 1000 simulations. "Reject(%)" denotes the percentage of the simulations where $H_0 : \theta = 0$ is rejected (i.e., type I error rate or power in this scenario), and "Time(m)" is measured in minutes.

Supplementary Table 3: Results summarized from 1000 simulations under the single exposure setting, with $K = 6$ total biomarkers. $\boldsymbol{\theta} = \mathbf{0/0.1}$, $H^2_{B_k} = \mathbf{0.2}$, and $H^2_x = 0.2$. The significance threshold for the IV selection is set at $\mathbf{5 \times 10^{-5}}$.

| $\theta$ | $N$ | $M$ | Method | Reject(%) | Time(m) |
|---|---|---|---|---|---|
| 0 | $6 \times 10^4$ | 18 | CaLMR (Uni) | 5.2 | 0.26 |
| | | | MRLE | 3.2 | 0.49 |
| | | | MR-PRESSO | 2.8 | 1.12 |
| | | | MR-IVW | 3.9 | - |
| | $8 \times 10^4$ | 35 | CaLMR (Uni) | 6.2 | 0.72 |
| | | | MRLE | 4.1 | 0.40 |
| | | | MR-PRESSO | 4.7 | 2.34 |
| | | | MR-IVW | 3.8 | - |
| | $1 \times 10^5$ | 63 | CaLMR (Uni) | 5.7 | 1.74 |
| | | | MRLE | 3.9 | 0.38 |
| | | | MR-PRESSO | 5.1 | 4.38 |
| | | | MR-IVW | 4.7 | - |
| 0.1 | $6 \times 10^4$ | 18 | CaLMR (Uni) | 43.8 | 0.22 |
| | | | MRLE | 41.7 | 0.46 |
| | | | MR-PRESSO | 2.9 | 1.15 |
| | | | MR-IVW | 3.9 | - |
| | $8 \times 10^4$ | 35 | CaLMR (Uni) | 86.2 | 0.44 |
| | | | MRLE | 84.5 | 0.46 |
| | | | MR-PRESSO | 5.1 | 2.34 |
| | | | MR-IVW | 3.9 | - |
| | $1 \times 10^5$ | 63 | CaLMR (Uni) | 98.6 | 1.75 |
| | | | MRLE | 98.1 | 0.40 |
| | | | MR-PRESSO | 8.6 | 4.46 |
| | | | MR-IVW | 8.1 | - |

"M" is the average number of IVs selected across 1000 simulations. "Reject(%)" denotes the percentage of the simulations where $H_0 : \theta = 0$ is rejected (i.e., type I error rate or power in this scenario), and "Time(m)" is measured in minutes.

Supplementary Table 4: Results summarized from 1000 simulations under the single exposure setting, with $K = 6$ total biomarkers. $\boldsymbol{\theta = 0/0.1}$, $H^2_{B_k} = \mathbf{0.3}$, and $H^2_x = 0.2$. The significance threshold for the IV selection is set at $\mathbf{5 \times 10^{-5}}$.

| $\theta$ | $N$ | $M$ | Method | Reject(%) | Time(m) |
|---|---|---|---|---|---|
| 0 | $6 \times 10^4$ | 48 | CaLMR (Uni) | 5.2 | 1.04 |
| | | | MRLE | 4.0 | 0.84 |
| | | | MR-PRESSO | 3.5 | 3.24 |
| | | | MR-IVW | 4.6 | - |
| | $8 \times 10^4$ | 95 | CaLMR (Uni) | 5.7 | 4.25 |
| | | | MRLE | 4.5 | 0.37 |
| | | | MR-PRESSO | 4.3 | 6.77 |
| | | | MR-IVW | 4.4 | - |
| | $1 \times 10^5$ | 148 | CaLMR (Uni) | 5.4 | 12.78 |
| | | | MRLE | 5.0 | 0.34 |
| | | | MR-PRESSO | 4.9 | 10.12 |
| | | | MR-IVW | 5.3 | - |
| 0.1 | $6 \times 10^4$ | 48 | CaLMR (Uni) | 95.4 | 0.87 |
| | | | MRLE | 94.9 | 0.39 |
| | | | MR-PRESSO | 6.1 | 3.01 |
| | | | MR-IVW | 6.3 | - |
| | $8 \times 10^4$ | 95 | CaLMR (Uni) | 100 | 3.54 |
| | | | MRLE | 100 | 0.40 |
| | | | MR-PRESSO | 8.5 | 5.99 |
| | | | MR-IVW | 8.9 | - |
| | $1 \times 10^5$ | 148 | CaLMR (Uni) | 100 | 11.38 |
| | | | MRLE | 100 | 0.41 |
| | | | MR-PRESSO | 14.9 | 9.45 |
| | | | MR-IVW | 16.8 | - |

"M" is the average number of IVs selected across 1000 simulations. "Reject(%)" denotes the percentage of the simulations where $H_0 : \theta = 0$ is rejected (i.e., type I error rate or power in this scenario), and "Time(m)" is measured in minutes.

#### 2.2 Simulation Results Under the Multiple Exposure Setting

In the multi-exposure simulations, we assume there are $L = 2$ total latent exposures underlying $K$ observable traits, and $cor(X_1, X_2) = -0.5$. Since MRLE required a minimum of $K = 2$ biomarkers co-regulated by the exposure, while $K = 3$ is near the borderline of convergence, we conducted simulations with a total of $K = 6/8$ biomarkers to check the model performance. We assume each exposure regulates three to five observed traits. As shown in the following Supplementary Tables, we observed that CaLMR can control the Type-I error rates sharply and maintain high powers. There were no convergence issues when applying CaLMR tests. The tables of the detailed simulation results are as follows.

Supplementary Table 5: Results summarized from 1000 simulations under the two exposure setting, with $K = 6/8$ total biomarkers. $\boldsymbol{(\theta_1, \theta_2) = (0, 0)}$, $cor(X_1, X_2) = -0.5$, and $\boldsymbol{K_{x_1} = K_{x_2} = K/2}$, i.e., there are no biomarkers regulated by two exposures simultaneously.

| $K$ | $N$ | $M$ | Method | $\text{RR}_{x_1}$(%) | $\text{RR}_{x_2}$(%) | Time(m) |
|---|---|---|---|---|---|---|
| 6 | $6 \times 10^4$ | 58 | CaLMR (Multi) | 4.7 | 5.2 | 2.56 |
| | | 36 — 38 | CaLMR (Uni) | 4.1 | 6.5 | 1.55 |
| | | 36 — 38 | MRLE | 4.8 | 5.9 | 0.32 |
| | | 58 | MVMR-PRESSO | 5.4 | 4.6 | 4.03 |
| | | 58 | MVMR-IVW | 5.3 | 5.2 | - |
| | $8 \times 10^4$ | 54 | CaLMR (Multi) | 4.9 | 4.8 | 2.42 |
| | | 32 — 34 | CaLMR (Uni) | 4.4 | 5.5 | 1.37 |
| | | 32 — 34 | MRLE | 4.2 | 5.1 | 0.37 |
| | | 54 | MVMR-PRESSO | 4.8 | 3.8 | 3.59 |
| | | 54 | MVMR-IVW | 4.9 | 4.4 | - |
| | $1 \times 10^5$ | 52 | CaLMR (Multi) | 4.3 | 5.6 | 2.34 |
| | | 30 — 31 | CaLMR (Uni) | 5.2 | 5.8 | 1.44 |
| | | 30 — 31 | MRLE | 4.9 | 5.5 | 0.21 |
| | | 52 | MVMR-PRESSO | 4.9 | 4.3 | 3.39 |
| | | 52 | MVMR-IVW | 5.1 | 4.6 | - |
| 8 | $6 \times 10^4$ | 79 | CaLMR (Multi) | 5.0 | 4.8 | 4.39 |
| | | 49 — 52 | CaLMR (Uni) | 6.1 | 4.4 | 2.49 |
| | | 49 — 52 | MRLE | 6.1 | 4.7 | 0.50 |
| | | 79 | MVMR-PRESSO | 4.9 | 5.4 | 6.27 |
| | | 79 | MVMR-IVW | 4.8 | 5.5 | - |
| | $8 \times 10^4$ | 72 | CaLMR (Multi) | 5.0 | 4.8 | 3.88 |
| | | 43 — 45 | CaLMR (Uni) | 5.7 | 4.6 | 2.09 |
| | | 43 — 45 | MRLE | 6.3 | 5.0 | 0.63 |
| | | 72 | MVMR-PRESSO | 5.4 | 5.5 | 5.47 |
| | | 72 | MVMR-IVW | 5.7 | 5.6 | - |
| | $1 \times 10^5$ | 69 | CaLMR (Multi) | 4.2 | 4.5 | 4.00 |
| | | 41 — 42 | CaLMR (Uni) | 5.0 | 4.6 | 2.25 |
| | | 41 — 42 | MRLE | 5.5 | 5.4 | 0.50 |
| | | 69 | MVMR-PRESSO | 5.6 | 4.7 | 5.19 |
| | | 69 | MVMR-IVW | 5.6 | 4.5 | - |

"M" is the average number of IVs selected across 1000 simulations. CaLMR (Uni) and MRLE contain two "M" values since they used exposure-specific IVs separately for each exposure. "$\text{RR}_{x_l}$(%)" denotes the percentage of the simulations where $H_0 : \theta_l = 0$ is rejected (i.e., type I error rate or power in this scenario), and "Time(m)" is measured in minutes.

Supplementary Table 6: Results summarized from 1000 simulations under the two exposure setting, with $K = 6/8$ total biomarkers. $\boldsymbol{(\theta_1, \theta_2) = (0, 0.1)}$, $cor(X_1, X_2) = -0.5$, and $\boldsymbol{K_{x_1} = K_{x_2} = K/2}$, i.e., there are no biomarkers regulated by two exposures simultaneously.

| $K$ | $N$ | $M$ | Method | $\text{RR}_{x_1}(\%)$ | $\text{RR}_{x_2}(\%)$ | Time(m) |
|---|---|---|---|---|---|---|
| | | 58 | CaLMR (Multi) | 4.2 | 44.1 | 2.58 |
| | | 36 — 38 | CaLMR (Uni) | 16.7 | 54.6 | 1.87 |
| | $6 \times 10^4$ | 36 — 38 | MRLE | 16.9 | 54.2 | 0.25 |
| | | 58 | MVMR-PRESSO | 5.3 | 9.8 | 3.59 |
| | | 58 | MVMR-IVW | 5.5 | 11.2 | - |
| | | 54 | CaLMR (Multi) | 4.8 | 64.8 | 2.54 |
| | | 32 — 34 | CaLMR (Uni) | 25.8 | 71.4 | 1.57 |
| 6 | $8 \times 10^4$ | 32 — 34 | MRLE | 23.4 | 69.2 | 0.18 |
| | | 54 | MVMR-PRESSO | 5.3 | 11.6 | 3.55 |
| | | 54 | MVMR-IVW | 5.5 | 12.6 | - |
| | | 52 | CaLMR (Multi) | 3.9 | 75.1 | 2.41 |
| | | 30 — 31 | CaLMR (Uni) | 32.3 | 80.4 | 1.44 |
| | $1 \times 10^5$ | 30 — 31 | MRLE | 31.1 | 78.4 | 0.16 |
| | | 52 | MVMR-PRESSO | 6.4 | 12.6 | 3.34 |
| | | 52 | MVMR-IVW | 6.6 | 14.4 | - |
| | | 79 | CaLMR (Multi) | 5.5 | 59.9 | 4.34 |
| | | 49 — 52 | CaLMR (Uni) | 24.7 | 66.6 | 3.03 |
| | $6 \times 10^4$ | 49 — 52 | MRLE | 22.4 | 66.3 | 0.49 |
| | | 79 | MVMR-PRESSO | 4.5 | 10.6 | 5.75 |
| | | 79 | MVMR-IVW | 4.8 | 11.4 | - |
| | | 72 | CaLMR (Multi) | 4.9 | 77.1 | 3.80 |
| | | 43 — 45 | CaLMR (Uni) | 33.1 | 82.9 | 2.44 |
| 8 | $8 \times 10^4$ | 43 — 45 | MRLE | 32.8 | 81.2 | 0.34 |
| | | 72 | MVMR-PRESSO | 5.7 | 10.5 | 5.49 |
| | | 72 | MVMR-IVW | 6.2 | 11.3 | - |
| | | 69 | CaLMR (Multi) | 4.0 | 87.5 | 3.63 |
| | | 41 — 42 | CaLMR (Uni) | 41.4 | 90.9 | 2.19 |
| | $1 \times 10^5$ | 41 — 42 | MRLE | 39.8 | 90.1 | 0.31 |
| | | 69 | MVMR-PRESSO | 5.4 | 12.1 | 5.14 |
| | | 69 | MVMR-IVW | 6.3 | 13.2 | - |

"M" is the average number of IVs selected across 1000 simulations. CaLMR (Uni) and MRLE contain two "M" values since they used exposure-specific IVs separately for each exposure. "$\text{RR}_{x_l}(\%)$" denotes the percentage of the simulations where $H_0 : \theta_l = 0$ is rejected (i.e., type I error rate or power in this scenario), and "Time(m)" is measured in minutes.

Supplementary Table 7: Results summarized from 1000 simulations under the two exposure setting, with $K = 6/8$ total biomarkers. $\boldsymbol{(\theta_1, \theta_2) = (0.1, 0.1)}$, $cor(X_1, X_2) = -0.5$, and $\boldsymbol{K_{x_1} = K_{x_2} = K/2}$, i.e., there are no biomarkers regulated by two exposures simultaneously.

| $K$ | $N$ | $M$ | Method | $\text{RR}_{x_1}$(%) | $\text{RR}_{x_2}$(%) | Time(m) |
|---|---|---|---|---|---|---|
| 6 | $6 \times 10^4$ | 58 | CaLMR (Multi) | 41.4 | 45.7 | 2.83 |
| | | 36 — 38 | CaLMR (Uni) | 19.6 | 41.0 | 1.57 |
| | | 36 — 38 | MRLE | 20.4 | 38.9 | 0.27 |
| | | 58 | MVMR-PRESSO | 8.5 | 10.0 | 4.06 |
| | | 58 | MVMR-IVW | 10.3 | 10.3 | - |
| | $8 \times 10^4$ | 54 | CaLMR (Multi) | 56.3 | 66.8 | 2.77 |
| | | 32 — 34 | CaLMR (Uni) | 25.9 | 62.4 | 1.35 |
| | | 32 — 34 | MRLE | 25.8 | 60.3 | 0.21 |
| | | 54 | MVMR-PRESSO | 10.6 | 10.2 | 3.82 |
| | | 54 | MVMR-IVW | 10.9 | 12.5 | - |
| | $1 \times 10^5$ | 52 | CaLMR (Multi) | 68.6 | 76.4 | 2.68 |
| | | 30 — 31 | CaLMR (Uni) | 32.0 | 72.8 | 1.27 |
| | | 30 — 31 | MRLE | 29.9 | 70.7 | 0.17 |
| | | 52 | MVMR-PRESSO | 12.5 | 11.5 | 3.55 |
| | | 52 | MVMR-IVW | 14.3 | 13.9 | - |
| 8 | $6 \times 10^4$ | 79 | CaLMR (Multi) | 50.7 | 61.8 | 5.10 |
| | | 49 — 52 | CaLMR (Uni) | 23.6 | 54.0 | 2.57 |
| | | 49 — 52 | MRLE | 22.7 | 54.0 | 0.52 |
| | | 79 | MVMR-PRESSO | 9.0 | 9.7 | 6.30 |
| | | 79 | MVMR-IVW | 10.4 | 10.5 | - |
| | $8 \times 10^4$ | 72 | CaLMR (Multi) | 68.4 | 77.9 | 3.62 |
| | | 43 — 45 | CaLMR (Uni) | 31.0 | 74.1 | 2.07 |
| | | 43 — 45 | MRLE | 30.7 | 71.8 | 0.39 |
| | | 72 | MVMR-PRESSO | 8.6 | 10.1 | 5.80 |
| | | 72 | MVMR-IVW | 10.3 | 11.2 | - |
| | $1 \times 10^5$ | 69 | CaLMR (Multi) | 78.2 | 88.4 | 3.57 |
| | | 41 — 42 | CaLMR (Uni) | 38.8 | 86.0 | 1.94 |
| | | 41 — 42 | MRLE | 37.6 | 83.8 | 0.31 |
| | | 69 | MVMR-PRESSO | 10.1 | 11.5 | 5.31 |
| | | 69 | MVMR-IVW | 11.1 | 13.2 | - |

"M" is the average number of IVs selected across 1000 simulations. CaLMR (Uni) and MRLE contain two "M" values since they used exposure-specific IVs separately for each exposure. "$\text{RR}_{x_l}$(%)" denotes the percentage of the simulations where $H_0 : \theta_l = 0$ is rejected (i.e., type I error rate or power in this scenario), and "Time(m)" is measured in minutes.

Supplementary Table 8: Results summarized from 1000 simulations with $K = 8$ total biomarkers and $\boldsymbol{K_{x_1}} = \boldsymbol{K_{x_2}} = \mathbf{5}$, i.e., there are two biomarkers regulated by two exposures simultaneously.

| $\theta_1$ | $\theta_2$ | $N$ | $M$ | Method | $\mathrm{RR}_{x_1}$(%) | $\mathrm{RR}_{x_2}$(%) | Time(m) |
|---|---|---|---|---|---|---|---|
| | | | 93 | CaLMR (Multi) | 4.0 | 4.5 | 4.81 |
| | | | 80 — 74 | CaLMR (Uni) | 4.6 | 4.7 | 6.25 |
| | | $6 \times 10^4$ | 80 — 74 | MRLE | 6.0 | 6.0 | 2.04* |
| | | | 93 | MVMR-PRESSO | 6.1 | 6.0 | 6.92 |
| | | | 93 | MVMR-IVW | 5.5 | 5.4 | - |
| | | | 90 | CaLMR (Multi) | 3.9 | 3.7 | 4.66 |
| | | | 78 — 70 | CaLMR (Uni) | 5.3 | 3.8 | 5.93 |
| 0 | 0 | $8 \times 10^4$ | 78 — 70 | MRLE | 6.4 | 7.5 | 2.08 |
| | | | 90 | MVMR-PRESSO | 4.2 | 4.1 | 6.74 |
| | | | 90 | MVMR-IVW | 4.2 | 4.0 | - |
| | | | 87 | CaLMR (Multi) | 4.3 | 3.8 | 4.35 |
| | | | 75 — 65 | CaLMR (Uni) | 5.0 | 4.5 | 5.36 |
| | | $1 \times 10^5$ | 75 — 65 | MRLE | 7.1 | 9.1 | 1.83 |
| | | | 87 | MVMR-PRESSO | 4.1 | 4.4 | 6.38 |
| | | | 87 | MVMR-IVW | 4.3 | 4.6 | - |
| | | | 93 | CaLMR (Multi) | 4.0 | 79.9 | 4.65 |
| | | | 80 — 74 | CaLMR (Uni) | 16.2 | 82.7 | 6.75 |
| | | $6 \times 10^4$ | 80 — 74 | MRLE | 35.6 | 80.3 | 2.93 |
| | | | 93 | MVMR-PRESSO | 6.7 | 11.3 | 6.90 |
| | | | 93 | MVMR-IVW | 8.0 | 12.1 | - |
| | | | 90 | CaLMR (Multi) | 4.6 | 93.7 | 4.64 |
| | | | 78 — 70 | CaLMR (Uni) | 24.1 | 95.6 | 5.72 |
| 0 | 0.1 | $8 \times 10^4$ | 78 — 70 | MRLE | 60.2 | 92.3 | 2.40 |
| | | | 90 | MVMR-PRESSO | 7.0 | 11.9 | 6.69 |
| | | | 90 | MVMR-IVW | 7.7 | 12.0 | - |
| | | | 93 | CaLMR (Multi) | 4.3 | 98.1 | 4.41 |
| | | | 80 — 74 | CaLMR (Uni) | 29.8 | 98.7 | 5.08 |
| | | $1 \times 10^5$ | 80 — 74 | MRLE | 74.3 | 95.8 | 2.08 |
| | | | 93 | MVMR-PRESSO | 6.7 | 11.1 | 6.44 |
| | | | 93 | MVMR-IVW | 7.4 | 11.5 | - |
| | | | 93 | CaLMR (Multi) | 50.2 | 80.1 | 4.62 |
| | | | 80 — 74 | CaLMR (Uni) | 47.6 | 79.6 | 6.44 |
| | | $6 \times 10^4$ | 80 — 74 | MRLE | 25.6 | 76.1 | 2.06 |
| | | | 93 | MVMR-PRESSO | 10.0 | 11.3 | 6.85 |
| | | | 93 | MVMR-IVW | 10.9 | 11.9 | - |
| | | | 90 | CaLMR (Multi) | 67.5 | 93.9 | 4.56 |
| | | | 78 — 70 | CaLMR (Uni) | 58.4 | 93.6 | 5.86 |
| 0.1 | 0.1 | $8 \times 10^4$ | 78 — 70 | MRLE | 35.3 | 90.5 | 2.19* |
| | | | 90 | MVMR-PRESSO | 9.7 | 11.8 | 6.94 |
| | | | 90 | MVMR-IVW | 10.8 | 12.4 | - |
| | | | 87 | CaLMR (Multi) | 79.7 | 98.4 | 4.31 |
| | | | 75 — 65 | CaLMR (Uni) | 66.7 | 98.2 | 5.26 |
| | | $1 \times 10^5$ | 75 — 65 | MRLE | 37.4 | 95.2 | 2.11* |
| | | | 87 | MVMR-PRESSO | 10.3 | 11.7 | 6.39 |
| | | | 87 | MVMR-IVW | 11.9 | 13.3 | - |

"M" is the average number of IVs selected across 1000 simulations. CaLMR (Uni) and MRLE contain two "M" values since they used exposure-specific IVs separately for each exposure. "$\mathrm{RR}_{x_l}$(%)" denotes the percentage of the simulations where $H_0 : \theta_l = 0$ is rejected (i.e., type I error rate or power in this scenario), and "Time(m)" is measured in minutes.

* MRLE had convergence issues in this setting.

### 3 Detailed Information on the Analysis of the Effects of Psychiatric Factors on Disease Risks

We applied our methods to test for the causal effects of 4 broad latent factors on a variety of potential disease outcomes. Grotzinger1 et al (2022) identified these four broad latent factors with their corresponding disorders by using genomic structural equation modeling. As stated in the Results section, these latent factors, derived from the genetic correlations observed among 11 psychiatric disorders, help explain both shared and unique genetic risks. Factor 1 defines general compulsive disorder and is characterized by the presence of specific disorders Anorexia Nervosa (AN), Obsessive Compulsive Disorder (OCD), and Tourette Syndrome (TS). Factor 2 defines general psychotic disorder and is characterized by the presence of specific disorders Schizophrenia (SCZ), Bipolar Disorder (BIP), and Alcohol Use Disorder (ALCH). Factor 3 defines general neurodevelopmental disorder and is characterized by the presence of specific disorders TS, ALCH, Attention-Deficit/Hyperactivity Disorder (ADHD), Autism Spectrum Disorder (AUT), Post-Traumatic Stress Disorder (PTSD), and Major Depressive Disorder (MDD). Finally, Factor 4 defines general internalizing disorder and is characterized by the presence of specific disorders ALCH, PTSD, MDD, and Generalized Anxiety Disorder (ANX). For our analysis, we treat the factors as latent exposures and use the set of specific psychiatric disorders associated with each factor as biomarkers. In total, there are $K = 11$ biomarkers across $L = 4$ latent factors with some biomarkers being associated with more than 1 latent factor (e.g. ALCH is associated with factors 2, 3, and 4) but this should not complicate the analysis.

Supplementary Table 9: Summary results for evaluating the causal effects of latent factor 1 (compulsive disorders) on 18 disease outcomes in the European population. Columns 2-6 list the p-values for each MR method, p-values are marked in bold if less than the overall adjusted significance level of $p < 0.0007$.

| Outcome | Original MRLE | MVMR-IVW | MVMR-PRESSO | CaLMR (Uni) | CaLMR (Multi) |
|---|---|---|---|---|---|
| ALS | 0.570 | 0.065 | 0.375 | 0.952 | 0.879 |
| Alzheimer's | $\mathbf{9.88 \times 10^{-21}}$ | 0.360 | 0.194 | 0.860 | 0.421 |
| Cardiac Arrhythmia | 1.000 | 0.089 | 0.388 | 0.946 | 0.415 |
| Cardiomyopathy | 0.971 | 0.053 | 0.628 | 0.995 | $\mathbf{3.75 \times 10^{-4}}$ |
| Celiac Disease | 1.000 | 0.037 | 0.421 | 0.967 | 0.070 |
| Crohn's Disease | 0.997 | 0.073 | 0.229 | 0.986 | 0.665 |
| Myasthenia Gravis | 0.136 | 0.066 | 0.504 | 0.946 | 0.712 |
| Heart Failure | 1.000 | 0.233 | 0.174 | 0.981 | 0.672 |
| IBS | 0.997 | 0.149 | 0.117 | 0.901 | 0.299 |
| Lupus | 0.985 | 0.241 | 0.027 | 0.906 | 0.996 |
| MS | 0.100 | 0.363 | 0.479 | 0.932 | 0.651 |
| Pancreatitis | 0.999 | 0.365 | 0.215 | 0.840 | 0.163 |
| Parkinson's | 1.000 | 0.065 | 0.270 | 0.950 | 0.180 |
| Psoriasis | 1.000 | 0.200 | 0.312 | 0.885 | 0.973 |
| RA | 1.000 | 0.579 | 0.288 | 0.961 | 0.957 |
| Type 1 Diabetes | 1.000 | 0.079 | 0.052 | 0.896 | 0.951 |
| Type 2 Diabetes | 0.601 | 0.047 | 0.141 | 0.871 | 0.715 |
| Vitiligo | 1.000 | 0.218 | 0.310 | 0.919 | 0.008 |

Supplementary Table 10: Summary results for evaluating the causal effects of latent factor 2 (psychotic disorders) on 18 disease outcomes in the European population. Columns 2-6 list the p-values for each MR method, p-values are marked in bold if less than the overall adjusted significance level of $p < 0.0007$.

| Outcome | Original MRLE | MVMR-IVW | MVMR-PRESSO | CaLMR (Uni) | CaLMR (Multi) |
|---|---|---|---|---|---|
| ALS | $\mathbf{6.23 \times 10^{-9}}$ | 0.417 | 0.347 | $\mathbf{1.23 \times 10^{-4}}$ | $\mathbf{4.23 \times 10^{-4}}$ |
| Alzheimer's | 0.021 | 0.598 | 0.180 | 0.916 | 0.001 |
| Cardiac Arrhythmia | 1.000 | 0.552 | 0.605 | 0.836 | $\mathbf{6.48 \times 10^{-4}}$ |
| Cardiomyopathy | 1.000 | 0.100 | 0.099 | 0.945 | $\mathbf{9.63 \times 10^{-5}}$ |
| Celiac Disease | 1.000 | 0.017 | 0.159 | 0.432 | $\mathbf{4.67 \times 10^{-10}}$ |
| Crohn's Disease | 0.962 | 0.013 | 0.524 | 0.876 | 0.328 |
| Myasthenia Gravis | 1.000 | 0.020 | 0.141 | $\mathbf{2.69 \times 10^{-8}}$ | 0.038 |
| Heart Failure | 1.000 | 0.158 | 0.365 | 0.997 | 0.013 |
| IBS | 1.000 | 0.286 | 0.687 | $\mathbf{4.68 \times 10^{-5}}$ | 0.013 |
| Lupus | $\mathbf{1.80 \times 10^{-20}}$ | 0.473 | 0.443 | $\mathbf{5.38 \times 10^{-11}}$ | $\mathbf{1.98 \times 10^{-6}}$ |
| MS | 0.904 | 0.025 | 0.351 | $\mathbf{1.54 \times 10^{-8}}$ | 0.001 |
| Pancreatitis | 1.000 | 0.099 | 0.087 | 0.984 | 0.035 |
| Parkinson's | 1.000 | 0.487 | 0.244 | $\mathbf{9.50 \times 10^{-4}}$ | 0.003 |
| Psoriasis | 0.153 | 0.106 | 0.017 | $\mathbf{2.06 \times 10^{-4}}$ | 0.140 |
| RA | 0.646 | 0.141 | 0.177 | 0.898 | 0.210 |
| Type 1 Diabetes | $\mathbf{4.84 \times 10^{-20}}$ | 0.095 | 0.318 | $\mathbf{3.55 \times 10^{-15}}$ | 0.330 |
| Type 2 Diabetes | 1.000 | 0.109 | 0.001 | $\mathbf{4.60 \times 10^{-8}}$ | 0.002 |
| Vitiligo | $\mathbf{9.16 \times 10^{-20}}$ | 0.782 | 0.532 | 0.919 | 0.005 |

Supplementary Table 11: Summary results for evaluating the causal effects of latent factor 3 (neurodevelopmental disorders) on 18 disease outcomes in the European population. Columns 2-6 list the p-values for each MR method, p-values are marked in bold if less than the overall adjusted significance level of $p < 0.0007$.

| Outcome | Original MRLE | MVMR-IVW | MVMR-PRESSO | CaLMR (Uni) | CaLMR (Multi) |
|---|---|---|---|---|---|
| ALS | $\mathbf{6.68 \times 10^{-21}}$ | $\mathbf{6.50 \times 10^{-4}}$ | 0.068 | $\mathbf{2.77 \times 10^{-11}}$ | 0.992 |
| Alzheimer's | $\mathbf{6.55 \times 10^{-21}}$ | 0.033 | 0.180 | $\mathbf{1.52 \times 10^{-4}}$ | 0.977 |
| Cardiac Arrhythmia | $\mathbf{4.20 \times 10^{-14}}$ | $9.03 \times 10^{-4}$ | 0.008 | 0.984 | 0.048 |
| Cardiomyopathy | 0.503 | 0.002 | 0.112 | 0.923 | 0.303 |
| Celiac Disease | $\mathbf{2.54 \times 10^{-21}}$ | 0.015 | 0.251 | 0.745 | $\mathbf{5.47 \times 10^{-4}}$ |
| Crohn's Disease | $\mathbf{1.11 \times 10^{-18}}$ | 0.212 | 0.054 | 0.937 | 0.742 |
| Myasthenia Gravis | 0.808 | 0.020 | 0.172 | 0.995 | 0.575 |
| Heart Failure | $\mathbf{6.68 \times 10^{-21}}$ | 0.016 | 0.091 | 0.957 | 0.204 |
| IBS | $\mathbf{1.80 \times 10^{-20}}$ | 0.033 | 0.591 | 0.920 | 0.068 |
| Lupus | $\mathbf{4.38 \times 10^{-13}}$ | 0.260 | 0.001 | 0.949 | $\mathbf{2.73 \times 10^{-12}}$ |
| MS | $\mathbf{2.45 \times 10^{-21}}$ | 0.425 | 0.239 | 0.972 | 0.976 |
| Pancreatitis | $\mathbf{6.55 \times 10^{-21}}$ | 0.092 | 0.111 | 0.027 | 0.092 |
| Parkinson's | $\mathbf{1.80 \times 10^{-20}}$ | 0.022 | 0.148 | 0.901 | 0.694 |
| Psoriasis | $\mathbf{1.80 \times 10^{-20}}$ | 0.072 | 0.036 | 0.734 | 0.990 |
| RA | $\mathbf{1.80 \times 10^{-20}}$ | 0.009 | 0.142 | 0.929 | 0.187 |
| Type 1 Diabetes | $\mathbf{6.55 \times 10^{-21}}$ | 0.051 | 0.036 | 0.203 | 0.766 |
| Type 2 Diabetes | $\mathbf{6.63 \times 10^{-21}}$ | 0.865 | $7.29 \times 10^{-4}$ | $\mathbf{2.22 \times 10^{-16}}$ | 0.939 |
| Vitiligo | $\mathbf{6.61 \times 10^{-21}}$ | 0.055 | 0.363 | 0.963 | 0.088 |

Supplementary Table 12: Summary results for evaluating the causal effects of latent factor 4 (internalizing disorders) on 18 disease outcomes in the European population. Columns list p-values for each MR method; p-values are marked in bold if less than the overall adjusted significance level of $p < 0.0007$.

| Outcome | Original MRLE | MVMR-IVW | MVMR-PRESSO | CaLMR (Uni) | CaLMR (Multi) |
|---|---|---|---|---|---|
| ALS | $\mathbf{1.58 \times 10^{-5}}$ | 0.002 | 0.068 | 0.869 | 0.955 |
| Alzheimer's | $\mathbf{2.42 \times 10^{-21}}$ | 0.146 | 0.180 | 0.970 | 0.954 |
| Cardiac Arrhythmia | $\mathbf{2.39 \times 10^{-21}}$ | 0.077 | 0.015 | 0.969 | 0.578 |
| Cardiomyopathy | $\mathbf{8.64 \times 10^{-15}}$ | 0.010 | 0.323 | 0.953 | 0.046 |
| Celiac Disease | $\mathbf{4.99 \times 10^{-7}}$ | 0.002 | 0.055 | $\mathbf{1.6 \times 10^{-30}}$ | 0.001 |
| Crohn's Disease | $\mathbf{2.39 \times 10^{-21}}$ | 0.484 | 0.260 | 0.996 | 0.093 |
| Myasthenia Gravis | $\mathbf{7.20 \times 10^{-21}}$ | 0.035 | 0.321 | 0.879 | 0.653 |
| Heart Failure | $\mathbf{3.25 \times 10^{-19}}$ | 0.052 | 0.365 | 0.985 | 0.155 |
| IBS | $\mathbf{7.92 \times 10^{-21}}$ | 0.082 | 0.393 | 0.969 | 0.236 |
| Lupus | 0.091 | 0.071 | 0.404 | 0.913 | $\mathbf{2.77 \times 10^{-30}}$ |
| MS | $\mathbf{2.38 \times 10^{-21}}$ | 0.142 | 0.351 | $\mathbf{6.32 \times 10^{-5}}$ | 0.863 |
| Pancreatitis | $\mathbf{1.81 \times 10^{-20}}$ | 0.378 | 0.087 | 0.871 | 0.265 |
| Parkinson's | 1.000 | 0.283 | 0.149 | 0.995 | 0.541 |
| Psoriasis | $\mathbf{1.89 \times 10^{-20}}$ | 0.250 | 0.036 | 0.798 | 0.778 |
| RA | $\mathbf{1.03 \times 10^{-13}}$ | 0.002 | 0.104 | 0.913 | 0.628 |
| Type 1 Diabetes | $\mathbf{3.60 \times 10^{-21}}$ | 0.451 | 0.036 | 0.817 | 0.972 |
| Type 2 Diabetes | $\mathbf{2.38 \times 10^{-21}}$ | 0.058 | 0.010 | 0.880 | 0.959 |
| Vitiligo | 1.000 | 0.306 | 0.167 | 0.964 | 0.077 |

### References

Jin Jin, Guanghao Qi, Zhi Yu, and Nilanjan Chatterjee. Mendelian randomization analysis using multiple biomarkers of an underlying common exposure. *Biostatistics*, page kxae006, 2024.
